# Artificial Intelligence Reveals Prognostic TP53 Pathway Alterations in FOLFOX-Treated Early-Onset Colorectal Cancer Among Populations at Risk

**DOI:** 10.1101/2025.10.27.25338923

**Authors:** Fernando C. Diaz, Brigette Waldrup, Francisco G. Carranza, Sophia Manjarrez, Enrique Velazquez-Villarreal

## Abstract

**Background:** The incidence of early-onset colorectal cancer (EOCRC; <50 years) continues to rise, with the most rapid increases observed among Hispanic/Latino (H/L) populations. Although the TP53 signaling pathway is a critical driver of colorectal tumorigenesis, its prognostic significance in FOLFOX-treated EOCRC—particularly among H/L patients— remains poorly defined.

**Methods:** We analyzed 2,515 colorectal cancer (CRC) cases (H/L = 266; non-Hispanic White [NHW] = 2,249), stratified by ancestry, age at onset, and FOLFOX treatment status. Statistical comparisons were performed using Fisher’s exact, chi-square, and Kaplan–Meier analyses. To enhance integrative data exploration, we applied AI-HOPE and AI-HOPE-TP53, novel conversational artificial intelligence (AI) platforms that enable natural language–driven analysis of multi-dimensional clinical, genomic, and therapeutic data.

**Results:** TP53 pathway alterations were detected in 85% of H/L and 83% of NHW patients treated with FOLFOX. Among late-onset (LO) cases, CHEK2 mutations were significantly lower in H/L patients compared with NHW (0.8% vs. 2.6%, *p* = 0.02), while TP53 mutations were more frequent in LO NHW patients receiving FOLFOX (78.9% vs. 73.4%, *p* = 0.04). Within LO NHW cases, FOLFOX-treated patients exhibited an enrichment of TP53 mutations (73.4% vs. 68.1%, *p* = 0.02) and lower frequencies of ATM (7.2% vs. 12.1%, *p* = 0.001) and CDKN2A (1.0% vs. 2.5%, *p* = 0.03) mutations compared to non-FOLFOX-treated counterparts. Across all cohorts, missense mutations represented the predominant alteration type. Remarkably, in EO H/L patients treated with FOLFOX, TP53 pathway alterations were associated with significantly improved overall survival (*p* = 0.014), an effect not observed in other subgroups.

**Conclusions:** TP53 pathway alterations may serve as an ancestry- and treatment-specific biomarker of favorable prognosis in FOLFOX-treated EOCRC H/L patients. The application of AI-driven integrative analytics through AI-HOPE platforms accelerated biomarker discovery and underscores the potential of conversational AI to advance precision oncology and health equity.

## 1. Introduction

Colorectal cancer (CRC) ranks as the third leading cause of both cancer-related deaths and incidence in the United States (1). Although widespread screening and prevention strategies have stabilized or reduced CRC rates in older adults, the disease remains a major public health challenge (2). Among individuals under 50 years of age—who often fall outside routine screening recommendations—CRC has become the leading cause of cancer-related deaths in men and the second in women (1). Alarmingly, by 2030, CRC is projected to become the leading cause of cancer-related mortality among individuals aged 20 to 49 (3–9). This increase in early-onset colorectal cancer (EOCRC) incidence is most pronounced among Hispanic/Latino (H/L) populations, who have experienced some of the steepest rises in both incidence and mortality in recent years (1, 2, 6, 10, 11). Representing 19% of the U.S. workforce and contributing approximately 15% (∼$4.2 trillion) to the U.S. GDP—a figure surpassing the economies of India, the United Kingdom, and France—the H/L population’s growing cancer burden poses an urgent public health and economic challenge (12). Understanding the molecular drivers of EOCRC in this disproportionately affected group is therefore essential for guiding prevention, therapy optimization, and equitable precision oncology initiatives.

Recent genomic studies have begun to uncover molecular distinctions between EOCRC and late-onset colorectal cancer (LOCRC). Unique features such as LINE-1 hypomethylation and distinct mutational profiles involving SMAD4, TP53, APC, and KRAS suggest that EOCRC may follow a biologically distinct trajectory (13–17). However, reported differences in tumor mutation burden, microsatellite instability (MSI), and PD-L1 expression remain inconsistent (15–17). Importantly, these investigations have largely focused on homogeneous, non-Hispanic White (NHW) cohorts, leaving significant gaps in the molecular characterization of EOCRC within underrepresented populations (18–20). This lack of diversity in genomic research contributes to persistent health disparities, as findings from one population are often extrapolated to others with differing genetic, environmental, and sociocultural contexts. Our group has recently addressed this gap by defining ancestry-specific alterations in key oncogenic pathways—including WNT, MAPK, JAK/STAT, PI3K, and TP53—in EOCRC among H/L patients (21–23). Yet, the functional and prognostic significance of these molecular features, particularly in the context of treatment response, remains poorly understood.

The TP53 tumor suppressor, often called the “guardian of the genome,” is the most frequently mutated gene across all cancer types (24). As the central regulator of the TP53 pathway, it orchestrates cell cycle arrest, apoptosis, DNA repair, and metabolic homeostasis (25, 26). In CRC, TP53 mutations occur in approximately 74% of tumors, with the majority being missense variants that confer altered or dominant-negative function (27). These mutations have been linked to chemoresistance and poor outcomes in various cancers (24). Within H/L CRC, our group previously observed higher rates of TP53 alterations compared to NHW patients, although no significant differences were found between early- and late-onset disease within the H/L subgroup (23). However, the impact of TP53 pathway alterations on treatment outcomes in this population remains unexplored.

For microsatellite-stable (MSS), mismatch repair–proficient (pMMR) metastatic CRC without actionable driver mutations, the ASCO-recommended first-line therapy consists of folinic acid, fluorouracil (5-FU), and oxaliplatin (FOLFOX) (25, 27). While this regimen is a cornerstone of CRC treatment, younger EOCRC patients experience higher toxicity and shorter overall survival compared to their late-onset counterparts (5). The role of TP53 in mediating FOLFOX response remains controversial: while some studies suggest improved sensitivity in tumors harboring wild-type TP53, others report no consistent correlation between TP53 status and treatment efficacy (28–30). Notably, these analyses have primarily examined European and Asian cohorts, leaving a critical void regarding H/L populations, who may harbor distinct mutational and pharmacogenomic profiles influencing drug response.

In this study, we conducted a comprehensive molecular and survival analysis of the TP53 pathway in early- and late-onset CRC among H/L patients treated with FOLFOX. We further applied AI-HOPE (31) and its specialized module AI-HOPE-TP53 (32), conversational artificial intelligence (AI) systems designed to integrate clinical, genomic, and treatment data through natural language–driven multi-parameter queries. This AI-enabled framework facilitates rapid, reproducible exploration of large-scale datasets, uncovering complex interactions between pathway alterations and clinical outcomes (33, 34). By leveraging this technology, we aim to define the prognostic significance of TP53 pathway alterations and their potential as ancestry- and treatment-specific biomarkers in FOLFOX-treated H/L EOCRC, advancing both precision oncology and health equity in cancer research.

## 2. Materials and Methods

### 2.1 Study design and data acquisition

This retrospective multi-cohort analysis integrated publicly available clinical and genomic data from CRC studies hosted on the cBioPortal for Cancer Genomics platform. The datasets included TCGA Colorectal Adenocarcinoma (PanCancer Atlas), MSK-CHORD, and AACR Project GENIE BPC CRC. These repositories were selected for their detailed annotation of somatic variants, treatment records, and survival outcomes. Only cases with histologically confirmed colorectal, colon, or rectal adenocarcinoma and corresponding primary tumor sequencing data were retained. For individuals represented by multiple tumor entries, a single sample per patient was randomly selected to prevent duplication bias.

### 2.2 Population stratification and demographic classification

Ancestry and ethnicity assignments followed a hierarchical approach prioritizing self-reported categories provided in the original datasets. Patients designated as “Hispanic or Latino,” “Spanish, NOS,” “Hispanic, NOS,” or “Latino, NOS” were classified as H/L. When explicit annotations were missing, surname-based algorithms validated for identifying Hispanic origin were applied. The comparator group included NHW patients meeting identical inclusion criteria. Age at diagnosis was derived from clinical metadata, and participants were grouped as early-onset CRC (EOCRC; <50 years) or late-onset CRC (LOCRC; ≥50 years).

### 2.3 Treatment annotation and FOLFOX classification

Therapy data were parsed to identify patients who had received the FOLFOX regimen, consisting of folinic acid (leucovorin), fluorouracil (5-FU), and oxaliplatin. To qualify as “FOLFOX-treated,” treatment timelines were verified to confirm overlapping or sequential administration of all three drugs, consistent with first-line standard-of-care protocols for metastatic, microsatellite-stable CRC. Patients lacking complete documentation of these agents were classified as non-FOLFOX-treated. Treatment records were further cross-checked against clinical notes and regimen codes to ensure internal consistency.

### 2.4 Annotation of TP53 pathway alterations

The TP53 signaling network was curated from peer-reviewed literature and canonical pathway databases, including genes directly involved in cell-cycle regulation, apoptosis, and DNA damage response. Somatic variants were retrieved from cBioPortal mutation calls and filtered to retain only non-synonymous alterations—including missense, nonsense, frameshift, splice-site, and start-loss variants. Pathway alteration status was defined by the presence of at least one qualifying mutation in any TP53 pathway gene. Variants of unknown significance and synonymous mutations were excluded to minimize false-positive associations.

### 2.5 Statistical and survival analysis

Differences in mutation frequencies between subgroups were evaluated using Fisher’s exact or chi-square tests, depending on expected cell counts. Continuous clinical variables were analyzed using the Mann–Whitney U test. Overall survival (OS) was calculated from the date of diagnosis to death or last follow-up. Survival distributions were estimated via the Kaplan–Meier method, with log-rank tests used to compare curves. Hazard ratios (HRs) and 95% confidence intervals (CIs) were generated using Cox proportional hazards regression, adjusted for age, sex, and ancestry. All analyses were performed in R (v4.3.2), and two-sided p values < 0.05 were considered statistically significant.

### 2.6 AI-HOPE-TP53 integration and automated analysis workflow

To streamline high-dimensional data interrogation, we implemented AI-HOPE-TP53, a specialized module within the AI-HOPE (Artificial Intelligence for Health Optimization and Precision Engagement) platform. This conversational AI system enables natural language–driven integration of clinical, genomic, and therapeutic data to execute complex multi-parameter queries and guide downstream analyses. Through iterative prompts such as “Among H/L patients treated with FOLFOX, do TP53 pathway alterations correlate with survival outcomes?” and “Compare mutation frequencies between EOCRC and LOCRC by ancestry and treatment group,” the AI system autonomously generated subgroup stratifications and mutation frequency matrices. Within this analytical framework, AI-HOPE-TP53 performed three interconnected tasks. First, it automatically identified eligible cohorts meeting combined criteria for ancestry, age at onset, treatment exposure, and TP53 pathway alteration status. Second, it generated detailed mutation landscapes, calculating pathway-specific alteration frequencies across defined subgroups. Finally, it conducted outcome association scans to prioritize variables demonstrating significant differences in survival or mutation patterns for formal statistical testing. This AI-assisted workflow minimized manual data handling, enhanced reproducibility, and accelerated the transition from exploratory analyses to validated hypothesis-driven results.

This conversational-AI–guided approach reduced manual preprocessing, improved reproducibility, and accelerated translation from exploratory interrogation to validated hypothesis testing, thereby bridging computational precision with biological insight.

## 3. Results

### 3.1 Baseline Clinical and Demographic Profiles of H/L and NHW CRC Cohorts

Table 1 summarizes the demographic and clinical characteristics of H/L and NHW CRC patients across a combined cohort of 2,515 individuals. Marked differences in age distribution were evident between groups: H/L patients were diagnosed, on average, 13.4 years earlier than NHW patients (47.8 ± 9.6 years vs. 61.2 ± 10.4 years; p < 0.001). Early-onset CRC (EOCRC; <50 years) accounted for 61% of all H/L cases but only 17% of NHW cases (p < 0.001), underscoring the disproportionate burden of early disease among H/L individuals.

**Table 1.** Overview of Hispanic/Latino (H/L) and Non-Hispanic White (NHW) colorectal cancer (CRC) cohorts, including age at diagnosis, FOLFOX treatment status, tumor characteristics, and ancestry.

| Clinical or Demographic Feature | H/L Cohort, n = 266 | NHW Cohort = 2,249 | Absolute Difference (% points) | p-value |
| --- | --- | --- | --- | --- |
| Age at Diagnosis (years, mean ± SD) | 47.8 ± 9.6 | 61.2 ± 10.4 | -13.4 | <0.001 |
| Early-Onset CRC (<50 years) | 162 (60.9%) | 374 (16.6%) | 44.3 | <0.001 |
| Late-Onset CRC (≥50 years) | 104 (39.1%) | 1,875 (83.4%) | -44.3 | |
| **Sex** |  |  |  |  |
| Male | 158 (59.4%) | 1,267 (56.3%) | 3.1 | 0.29 |
| Female | 108 (40.6%) | 982 (43.7%) | -3.1 |  |
| **FOLFOX Treatment Status** |  |  |  |  |
| Treated with FOLFOX | 164 (61.6%) | 1,294 (57.6%) | 4 | 0.17 |
| Not Treated with FOLFOX | 102 (38.3%) | 955 (42.4%) | -4.1 |  |
| **Stage at Diagnosis** |  |  |  |  |
| Stage I-III (Localized/Regional) | 156 (58.6%) | 1,236 (55.0%) | 3.6 | 0.28 |
| Stage IV (Metastatic) | 108 (40.6%) | 1,005 (44.7%) | -4.1 |  |
| NA | 2 (0.8%) | 8 (0.4%) | 0.4 |  |
| **Microsatellite Instability (MSI) Status** |  |  |  |  |
| MSI-Stable (MSS) | 200 (75.2%) | 1,940 (86.3%) | -11.1 | <0.001 |
| MSI-High / Instable | 21 (7.9%) | 209 (9.3%) | -1.4 | 0.46 |
| Indeterminate | 10 (3.8%) | 57 (2.5%) | 1.3 |  |
| NA | 35 (13.2%) | 43 (1.9%) | 11.3 |  |
| **Ethnic Subclassification (H/L only)** |  |  |  |  |
| Mexican / Chicano | 30 (11.3%) | — | — | — |
| Spanish, Hispanic, Latino, NOS | 236 (88.7%) | — | — | — |
| Non-Hispanic White | — | 2,249 (100.0%) | — | — |
| FOLFOX-Treated EOCRC Only |  |  |  |  |
| TP53 Pathway Altered | 140 (85.4%) | 1,076 (83.2%) | 2.2 | 0.34 |
| Wild-Type / Not Altered | 24 (14.6%) | 218 (16.8%) | -2.2 |  |

Sex distribution was similar across populations, with a modest male predominance in both groups (59.4% in H/L vs. 56.3% in NHW, p = 0.29). Receipt of FOLFOX therapy was also comparable, with 61.6% of H/L and 57.6% of NHW patients receiving the regimen (p = 0.17). When stratified by disease stage, localized or regional disease (Stage I–III) was slightly more frequent among H/L patients (58.6% vs. 55.0%), whereas advanced (Stage IV) disease occurred marginally less often (40.6% vs. 44.7%), though neither difference reached statistical significance.

Microsatellite instability (MSI) profiling revealed notable contrasts: MSI-stable (MSS) tumors predominated in both cohorts but were less common among H/L patients (75.2% vs. 86.3%; p < 0.001). The H/L cohort also exhibited a higher proportion of missing MSI data (13.2% vs. 1.9%), which may reflect underreporting or incomplete testing in this population. The frequency of MSI-high and indeterminate cases did not differ significantly between groups.

Ethnic subclassification confirmed distinct ancestry assignments. All NHW cases were classified as non-Hispanic/non-Spanish, while the H/L cohort was composed primarily of patients annotated as “Spanish, Hispanic, or Latino, NOS” (88.7%), followed by those identified as “Mexican/Chicano” (11.3%).

Among FOLFOX-treated EOCRC patients, TP53 pathway alterations were highly prevalent in both ancestries (85.4% in H/L vs. 83.2% in NHW; p = 0.34), suggesting that pathway disruption is common irrespective of ethnic background. Collectively, these data highlight substantial ancestry- and age-related disparities in CRC presentation, with H/L patients exhibiting a younger age of onset, lower MSI-stable frequency, and comparable TP53 alteration rates despite treatment similarities.

### 3.2 Comparative Genomic Analysis by Age and Ancestry

#### Hispanic/Latino Patients Stratified by Age and FOLFOX Treatment

Table 2a presents clinical and genomic characteristics of H/L CRC patients, analyzed according to age at diagnosis and FOLFOX treatment status. Among early-onset cases, the median age at diagnosis was slightly higher in FOLFOX-treated patients (42 years; IQR 36–47) compared with those who did not receive FOLFOX (40 years; IQR 34–43), with a near-significant trend (p = 0.0541). In contrast, among late-onset patients, those treated with FOLFOX were diagnosed at a significantly younger age (59 years; IQR 54–66) relative to non-treated counterparts (62 years; IQR 56–70; p = 0.0487). Across both early- and late-onset groups, mutation counts were broadly similar between treatment arms, with no significant differences observed. In EOCRC H/L patients, the TMB showed a modest, non-significant tendency toward higher values in the non-FOLFOX subgroup. However, in late-onset disease, non-FOLFOX-treated patients displayed a significantly elevated TMB (6.9; IQR 5.6–9.0) compared with FOLFOX-treated individuals (6.1; IQR 4.9–7.8; p = 0.0439).

**Table 2.** Clinical and Genomic Comparison of Early-Onset and Late-Onset Colorectal Cancer (CRC) Cohorts. It presents an integrated overview of clinical and molecular features distinguishing early-onset (EOCRC) and late-onset (LOCRC) colorectal cancer across major patient subgroups. The analysis encompasses four principal comparisons: (A) EOCRC versus LOCRC among Hispanic/Latino (H/L) patients; (B) EOCRC versus LOCRC among Non-Hispanic White (NHW) patients; (C) cross-ethnic comparison of EOCRC between H/L and NHW cohorts; and (D) stratified evaluation of EOCRC and LOCRC in H/L patients by FOLFOX treatment exposure. Key parameters include median age at diagnosis, overall somatic mutation burden, and the frequency of alterations in TP53 pathway genes, analyzed within each ancestry and age-defined subgroup. This comparative framework highlights the interplay between genomic disruption, treatment status, and demographic factors, providing insight into how early disease onset may shape the molecular landscape of CRC across ethnically diverse populations.

| Clinical Feature | Early-Onset Hispanic/Latino Treated with FOLFOX n (%) | Early-Onset Hispanic/Latino Not Treated with FOLFOX n (%) | p-value | Late-Onset Hispanic/Latino Treated with FOLFOX n (%) | Late-Onset Hispanic/Latino Not Treated with FOLFOX n (%) | p-value |
| --- | --- | --- | --- | --- | --- | --- |
| Median Diagnosis Age (IQR) | 42 (36-47) | 40 (34-43) | 0.05411 | 59 (54-66) | 62 (56-70) | **0.04865** |
| Median Mutation Count | 7 (5-8) | 7 (5-20) | 0.09735 | 8 (6-9) [NA=1] | 7 (5.25-9) | 0.6507 |
| Median TMB (IQR) | 6.3 (4.5-7.8) [NA=15] | 5.5 (3.4-8.3) [NA=2] | 0.1719 | 6.1 (4.9-7.8) [NA=10] | 6.9 (5.6-9.0) [NA=2] | 0.04389 |
| Median FGA | 0.18 (0.03-0.27) [NA=6] | 0.19 (0.03-0.29) | 0.7661 | 0.15 (0.06-0.25) [NA=7] | 0.21 (0.04-0.3) [NA=2] | 0.5464 |
| **TP53 Mutation** |  |  |  |  |  |  |
| Present | 57 (78.1%) | 42 (80.8%) | 0.8876 | 71 (78.0%) | 36 (72.0%) | 0.5526 |
| Absent | 16 (21.9%) | 10 (19.2%) |  | 20 (22.0%) | 14 (28.0%) |  |

(B)
| Clinical Feature | Early-Onset NHW<br>Treated with FOLFOX<br>n (%) | Early-Onset NHW<br>Not Treated with<br>FOLFOX<br>n (%) | p-value | Late-Onset NHW<br>Treated with FOLFOX<br>n (%) | Late-Onset NHW<br>Not Treated with<br>FOLFOX<br>n (%) | p-value |
| --- | --- | --- | --- | --- | --- | --- |
| Median Diagnosis Age (IQR) | 43 (37-48) | 44 (38-47) | 0.5646 | 63 (57-69) | 66 (57-74) | 4.146E-07 |
| Median Mutation Count | 6 (5-8) [NA=4] | 7 (5-9) [NA=2] | 0.1258 | 7 (5-9) [NA=10] | 8 (6-12) [NA=3] | 1.22E-05 |
| Median TMB (IQR) | 5.7 (4.1-6.9) | 5.7 (4.1-7.8) | 0.4214 | 6.1 (4.3-8.2) | 6.6 (4.9-10.4) | 0.0002854 |
| Median FGA | 0.14 (0.04-0.24) [NA=4] | 0.15 (0.04-0.23) [NA=2] | 0.5589 | 0.16 (0.06-0.28) [NA=6] | 0.15 (0.05-0.27) [NA=5] | 0.1929 |
| TP53 Mutation |  |  |  |  |  |  |
| Present | 296 (78.9%) | 228 (75.5%) | 0.3319 | 675 (73.4%) | 445 (68.1%) | 0.02559 |
| Absent | 79 (21.1%) | 74 (24.5%) |  | 244 (26.6%) | 208 (31.9%) |  |
| CDKN2A Mutation |  |  |  |  |  |  |
| Present | 3 (0.8%) | 5 (1.7%) | 0.4772 | 9 (1.0%) | 16 (2.5%) | 0.03638 |
| Absent | 372 (99.2%) | 297 (98.3%) |  | 910 (99.0%) | 637 (97.5%) |  |
| ATM Mutation |  |  |  |  |  |  |
| Present | 19 (5.1%) | 24 (7.9%) | 0.171 | 66 (7.2%) | 79 (12.1%) | 0.001233 |
| Absent | 356 (94.9%) | 278 (92.1%) |  | 853 (92.8%) | 574 (87.9%) |  |

(C)
| Clinical Feature | Early-Onset Hispanic/Latino Treated with FOLFOX n (%) | Early-Onset NHW Treated with FOLFOX n (%) | p-value | Early-Onset Hispanic/Latino Not Treated with FOLFOX n (%) | Early-Onset NHW Not Treated with FOLFOX n (%) | p-value |
| --- | --- | --- | --- | --- | --- | --- |
| Median Diagnosis Age (IQR) | 42 (36-47) | 43 (37-48) | 0.08467 | 40 (34-43) | 44 (38-47) | 0.0006016 |
| Median Mutation Count | 7 (5-8) | 6 (5-8) [NA=4] | 0.942 | 7 (5-20) | 7 (5-9) [NA=2] | 0.2601 |
| Median TMB (IQR) | 6.3 (4.5-7.8) [NA=15] | 5.7 (4.1-6.9) | 0.05806 | 5.5 (3.4-8.3) [NA=2] | 5.7 (4.1-7.8) | 0.5732 |
| Median FGA | 0.18 (0.03-0.27) [NA=6] | 0.14 (0.04-0.24) [NA=4] | 0.5556 | 0.19 (0.03-0.29) | 0.15 (0.04-0.23) [NA=2] | 0.3612 |
| ATM Mutation |  |  |  |  |  |  |
| Present | 5 (6.8%) | 19 (5.1%) | 0.7378 | 9 (17.3%) | 24 (7.9%) | 0.05927 |
| Absent | 68 (93.2%) | 356 (94.9%) |  | 43 (82.7%) | 278 (92.1%) |  |
| TSC1 Mutation |  |  |  |  |  |  |
| Present | 2 (2.7%) | 7 (1.9%) | 0.6445 | 6 (11.5%) | 10 (3.3%) | 0.02282 |
| Absent | 71 (97.3%) | 368 (98.1%) |  | 46 (88.5%) | 292 (96.7%) |  |
| RPTOR Mutation |  |  |  |  |  |  |
| Present | 2 (2.7%) | 10 (2.7%) | 1 | 4 (7.7%) | 6 (2.0%) | 0.04426 |
| Absent | 71 (97.3%) | 365 (97.3%) |  | 48 (92.3%) | 296 (98.0%) |  |

The fraction of genome altered (FGA) remained comparable across all age and treatment categories, suggesting that global genomic instability was not substantially influenced by FOLFOX exposure. TP53 mutations were prevalent in both early- and late-onset cohorts, detected in approximately 78–81% of cases, with no statistically significant associations observed between mutation status and treatment group. These findings indicate that while FOLFOX treatment correlates with subtle age and TMB differences, the overall mutation burden, genomic alteration patterns, and TP53 status appear largely consistent across subgroups of H/L patients.

#### Non-Hispanic White Patients Stratified by Age and FOLFOX Treatment

Table 2b summarizes the clinical and molecular characteristics of NHW CRC patients, stratified by both age group and FOLFOX treatment status. Among early-onset cases, age at diagnosis was comparable between those treated with FOLFOX and those who were not (43 years vs. 44 years; p = 0.56), indicating no treatment-related difference in diagnostic timing. However, in the late-onset population, FOLFOX-treated patients were diagnosed at a significantly younger age (63 years; IQR 57–69) compared with their non-treated counterparts (66 years; IQR 57–74; p = 4.15 × 10⁻⁷).

Genomic metrics revealed age- and treatment-dependent patterns. Mutation burden did not differ significantly among early-onset patients, but in late-onset cases, non-FOLFOX patients exhibited a higher median mutation count (8; IQR 6–12) than those receiving FOLFOX (7; IQR 5–9; p = 1.22 × 10⁻⁵). A similar trend was observed for TMB, which remained stable in early-onset disease yet rose significantly among late-onset non-treated patients (6.6 vs. 6.1; p = 2.85 × 10⁻⁴). The FGA was consistent across all groups, showing no association with treatment exposure.

At the gene-specific level, several key differences emerged. TP53 mutations were highly prevalent across all NHW subgroups but occurred less frequently in late-onset FOLFOX-treated patients (73.4%) compared with those not treated (68.1%; p = 0.026). Among DNA damage repair genes, ATM mutations were more frequent in late-onset non-FOLFOX patients (12.1%) than in those receiving FOLFOX (7.2%; p = 0.0012). Similarly, CDKN2A alterations were enriched in non-treated late-onset cases (2.5% vs. 1.0%; p = 0.036). These findings suggest that while early-onset NHW CRCs exhibit relatively stable genomic profiles regardless of therapy, late-onset tumors not treated with FOLFOX show greater mutational complexity, including higher mutation counts and a greater prevalence of alterations in TP53, ATM, and CDKN2A.

#### Ethnic Differences in Early-Onset Colorectal Cancer

Table 2c summarizes EOCRC characteristics in H/L and NHW patients, separated by FOLFOX treatment status. Among patients who received FOLFOX, H/L cases were diagnosed at a slightly younger age than NHW patients (42 vs. 43 years), a difference that did not reach statistical significance (p = 0.085). However, in the non-FOLFOX group, the age gap widened considerably, with H/L patients diagnosed significantly earlier (40 vs. 44 years; p = 0.0006).

Across both treatment strata, mutation counts and TMB were comparable, although a modest upward trend in TMB was observed among FOLFOX-treated H/L patients (median 6.3 vs. 5.7; p = 0.058). The FGA remained consistent between ethnicities, suggesting similar overall genomic instability.

Notable differences emerged in specific gene alterations. TSC1 mutations occurred more frequently in non-FOLFOX-treated H/L patients (11.5%) than in NHW patients (3.3%; p = 0.0228), while RPTOR mutations showed a comparable enrichment in the same subgroup (7.7% vs. 2.0%; p = 0.0443). Conversely, ATM mutation rates were similar between treated cohorts but tended to be higher among non-FOLFOX H/L patients (17.3% vs. 7.9%; p = 0.059).

Collectively, these findings indicate that while overall mutation load and genomic alteration profiles were largely comparable between early-onset H/L and NHW cases, H/L patients tended to present at younger ages and showed increased frequencies of TSC1 and RPTOR mutations, particularly in the absence of FOLFOX therapy.

### 3.3 TP53-Beta Pathway Alterations by Age, Ancestry, and Treatment Status

TP53 pathway alteration patterns were analyzed in H/L and NHW CRC cohorts according to patient age at diagnosis and FOLFOX therapy exposure (Tables 3a–3d).

**Table 3.** TP53 Pathway Alterations Across Age, Ancestry, and Treatment Subgroups in Colorectal Cancer. It presents the distribution of mutations affecting the TP53 signaling pathway among colorectal cancer (CRC) patients, analyzed according to age of onset, ancestry, and FOLFOX treatment status. The analysis is divided into four comparative frameworks: (3a) early-onset (EOCRC) versus late-onset (LOCRC) within the Hispanic/Latino (H/L) cohort, further separated by treatment exposure; (3b) FOLFOX-treated versus untreated subgroups within EOCRC and LOCRC among Non-Hispanic White (NHW) patients; (3c) comparison of EOCRC cases between H/L and NHW populations by treatment status; and (3d) analogous comparison for LOCRC patients. The table details mutation frequencies of major genes implicated in TP53 pathway regulation and evaluates their variation across these clinical contexts. This stratified approach captures potential interactions among age, ancestry, and chemotherapy exposure, revealing how these factors jointly influence TP53 pathway dysregulation and molecular heterogeneity in CRC.

| Pathway Alterations | Early-Onset<br>Hispanic/Latino<br>Treated with<br>FOLFOX<br>n (%) | Early-Onset<br>Hispanic/Latino<br>Not Treated<br>with FOLFOX<br>n (%) | p-value | Late-Onset<br>Hispanic/Latino<br>Treated with<br>FOLFOX<br>n (%) | Late-Onset<br>Hispanic/Latino<br>Not Treated<br>with FOLFOX<br>n (%) | p-value |
| --- | --- | --- | --- | --- | --- | --- |
| TP53 Alterations Present | 62 (84.9%) | 48 (92.3%) | 0.2702 | 78 (85.7%) | 38 (76.0%) | 0.2246 |
| TP53 Alterations Absent | 11 (15.1%) | 4 (7.7%) |  | 13 (14.3%) | 12 (24.0%) |  |

(B)
| Pathway Alterations | Early-Onset<br>NHW<br>Treated with<br>FOLFOX<br>n (%) | Early-Onset<br>NHW<br>Not Treated<br>with FOLFOX<br>n (%) | p-value | Late-Onset<br>NHW<br>Treated with<br>FOLFOX<br>n (%) | Late-Onset<br>NHW<br>Not Treated<br>with FOLFOX<br>n (%) | p-value |
| --- | --- | --- | --- | --- | --- | --- |
| TP53 Alterations Present | 311 (82.9%) | 247 (81.8%) | 0.7737 | 718 (78.1%) | 499 (76.4%) | 0.4601 |
| TP53 Alterations Absent | 64 (17.1%) | 55 (18.2%) |  | 201 (21.9%) | 154 (23.6%) |  |

(C)
| Pathway Alterations | Early-Onset<br>Hispanic/Latino<br>Treated with<br>FOLFOX<br>n (%) | Early-Onset<br>NHW<br>Treated<br>with<br>FOLFOX<br>n (%) | p-value | Early-Onset<br>Hispanic/Latino<br>Not Treated<br>with FOLFOX<br>n (%) | Early-Onset<br>NHW<br>Not Treated<br>with<br>FOLFOX<br>n (%) | p-value |
| --- | --- | --- | --- | --- | --- | --- |
| TP53 Alterations Present | 62 (84.9%) | 311 (82.9%) | 0.8049 | 48 (92.3%) | 247 (81.8%) | 0.06949 |
| TP53 Alterations Absent | 11 (15.1%) | 64 (17.1%) |  | 4 (7.7%) | 55 (18.2%) |  |

(D)
| Pathway Alterations | Late-Onset<br>Hispanic/Latino<br>Treated with<br>FOLFOX<br>n (%) | Late-Onset<br>NHW<br>Treated<br>with<br>FOLFOX<br>n (%) | p-value | Late-Onset<br>Hispanic/Latino<br>Not Treated<br>with FOLFOX<br>n (%) | Late-Onset<br>NHW<br>Not Treated<br>with<br>FOLFOX<br>n (%) | p-value |
| --- | --- | --- | --- | --- | --- | --- |
| TP53 Alterations Present | 78 (85.7%) | 718 (78.1%) | 0.12 | 38 (76.0%) | 499 (76.4%) | 1 |
| TP53 Alterations Absent | 13 (14.3%) | 201 (21.9%) |  | 12 (24.0%) | 154 (23.6%) |  |

Within the H/L cohort, the prevalence of TP53 pathway alterations remained broadly consistent across treatment groups in both EOCRC and LOCRC. Among early-onset patients, alterations were detected in 84.9% of those receiving FOLFOX and 92.3% of those who did not (p = 0.27), indicating no significant association between chemotherapy exposure and mutation frequency. In the late-onset subgroup, TP53 alterations were similarly frequent, observed in 85.7% of FOLFOX-treated cases compared with 76.0% in untreated patients (p = 0.22). These findings suggest that TP53 pathway disruption is highly prevalent and largely independent of treatment status across both age groups in the H/L population.

Among NHW patients, the frequency of TP53 pathway alterations was comparable across treatment groups in both EOCRC and LOCRC. In the early-onset subgroup, TP53 alterations were identified in 82.9% of FOLFOX-treated cases and 81.8% of those who did not receive FOLFOX (p = 0.77), indicating no meaningful treatment-related difference. Similarly, in late-onset NHW patients, alterations occurred in 78.1% of treated and 76.4% of untreated cases (p = 0.46). These data suggest that TP53 pathway mutations are highly prevalent across NHW CRC patients and appear largely unaffected by FOLFOX exposure or age of onset.

When comparing EOCRC cases across ancestries, TP53 pathway alterations occurred at similar frequencies between H/L and NHW patients. Among those who received FOLFOX, 84.9% of H/L patients and 82.9% of NHW patients exhibited TP53 alterations, with no significant difference detected (p = 0.80). In the non-FOLFOX subgroup, the prevalence of TP53 alterations was somewhat higher among H/L patients (92.3%) compared to NHW patients (81.8%), a difference approaching borderline statistical significance (p = 0.06). Overall, these results indicate that ancestry-related differences in TP53 alteration frequency are minimal in EOCRC, regardless of treatment exposure.

In LOCRC, TP53 pathway alterations occurred at comparable frequencies across ancestries, with only modest variation by treatment status. Among patients who received FOLFOX, H/L individuals exhibited a higher alteration rate (85.7%) relative to NHW patients (78.1%), though this difference did not reach statistical significance (p = 0.12). Similarly, in the non-FOLFOX subgroup, alteration prevalence remained nearly identical between H/L (76.0%) and NHW (76.4%) patients (p = 1.00). These results suggest that ancestry exerts minimal influence on TP53 mutation frequency in late-onset disease, regardless of FOLFOX treatment exposure.

### 3.4 Frequencies of Gene Alterations in the TGF-Beta Pathway

#### Early-Onset Hispanic/Latino Patients

Table S1 summarizes the distribution of TP53 pathway gene mutations in EOCRC H/L patients according to FOLFOX treatment status. TP53 mutations were highly prevalent across both groups—identified in 78.1% of FOLFOX-treated cases and 80.8% of non-treated cases (p = 0.89)—indicating no treatment-related difference. Mutations in CDKN2A and CHEK2 were rare, each observed in fewer than 2% of samples, while MDM2 and MDM4 alterations were absent entirely. A modest enrichment of ATM mutations was noted among patients who did not receive FOLFOX (17.3%) compared with those treated (6.8%), though this trend did not reach statistical significance (p = 0.12). Overall, the data suggest that TP53 pathway disruption is widespread, driven primarily by TP53 itself, with minimal contribution from other regulators such as MDM2, MDM4, or CDKN2A.

#### Late-Onset Hispanic/Latino Patients

As shown in Table S2, TP53 mutations were the most frequent alterations detected among LOCRC H/L patients, occurring in 78.0% of FOLFOX-treated cases and 72.0% of non-treated cases (p = 0.55). Other TP53 pathway components were rarely affected. MDM2 mutations were observed only in the treated group (2.2%), while MDM4 alterations were completely absent across both subgroups. Similarly, CDKN2A and CHEK2 mutations were infrequent, each identified in ≤1% of cases. ATM mutations were slightly more common in untreated patients (12.0%) compared with those receiving FOLFOX (8.8%), though this difference was not statistically significant (p = 0.75).

#### Early-Onset Non-Hispanic White Patients

Table S3 summarizes TP53 pathway mutation frequencies among EOCRC NHW patients stratified by FOLFOX treatment status. TP53 mutations were the most frequent alteration, detected in 78.9% of FOLFOX-treated patients and 75.5% of those who did not receive chemotherapy (p = 0.33), showing no significant treatment-related difference. Mutations in ATM and CHEK2 were relatively uncommon, present in 5–8% and 1–3% of cases, respectively, while CDKN2A mutations were observed in fewer than 2% of patients. Alterations in MDM2 and MDM4 were rare, occurring in less than 2% of cases overall.

#### Late-Onset Non-Hispanic White Patients

Table S4 summarizes mutation frequencies in TP53 pathway genes among LOCRC NHW patients, stratified by FOLFOX treatment status. TP53 mutations were the most common event, occurring in 73.4% of FOLFOX-treated and 68.1% of untreated patients—a statistically significant difference (p = 0.026) suggesting a modest enrichment among treated cases. Mutations in other pathway regulators were relatively uncommon. ATM alterations were observed more frequently in untreated patients (12.1%) than in those who received FOLFOX (7.2%; p = 0.0012), while CDKN2A mutations followed a similar pattern (2.5% vs. 1.0%; p = 0.036).

#### Comparison of Early- and Late-Onset Hispanic/Latino Patients Treated with FOLFOX

Table S5 presents the mutational profiles of TP53 pathway genes among H/L CRC patients, stratified by age at diagnosis. TP53 mutations were detected at nearly identical rates in early-onset (78.1%) and late-onset (78.0%) FOLFOX-treated patients (p = 1.00), underscoring the stability of TP53 alteration prevalence across age groups. Mutations in MDM2, CDKN2A, ATM, and CHEK2 occurred infrequently, each affecting fewer than 10% of cases and showing no significant age-related differences. MDM4 alterations were entirely absent across both subgroups.

#### Comparison of Early- and Late-Onset Hispanic/Latino Patients Not Treated with FOLFOX

Table S6 details the distribution of TP53 pathway mutations among untreated H/L CRC patients, stratified by age of onset. TP53 mutations were highly recurrent in both groups, occurring in 80.8% of early-onset and 72.0% of late-onset patients (p = 0.42), indicating no statistically significant age-related difference. Other pathway components showed minimal mutational activity. ATM alterations were among the more frequent secondary events, detected in 17.3% of early-onset and 12.0% of late-onset patients, though the difference was not significant (p = 0.63).

#### Comparison of Early- and Late-Onset Non-Hispanic White Patients Treated with FOLFOX

Table S7 summarizes TP53 pathway mutation frequencies in FOLFOX-treated NHW CRC patients, comparing EOCRC and LOCRC groups. TP53 mutations were widespread in both cohorts but appeared slightly more frequent in early-onset patients (78.9%) than in late-onset cases (73.4%), a modest yet statistically significant difference (*p* = 0.046). Alterations in other TP53 regulatory genes were uncommon. ATM and CHEK2 mutations occurred in fewer than 8% and 2% of cases, respectively, while MDM2, MDM4, and CDKN2A alterations were detected in less than 1% of patients overall. A small increase in MDM2 mutations was observed in early-onset individuals (1.1%) compared with late-onset patients (0.2%), though this did not reach statistical significance (*p* = 0.06*).

#### Early-Onset FOLFOX-Treated Patients: Hispanic/Latino vs. Non-Hispanic White Comparisons

Table S8 compares the mutational landscape of TP53 pathway genes between EOCRC H/L and NHW patients treated with FOLFOX. TP53 mutations were highly prevalent and nearly identical in frequency between the two groups (78.1% in H/L vs. 78.9% in NHW; p = 1.00), underscoring the shared centrality of TP53 pathway disruption across ancestries. Alterations in ATM, CHEK2, and CDKN2A were uncommon and exhibited comparable distributions across both cohorts. MDM2 and MDM4 mutations were exceedingly rare, detected only in NHW patients at frequencies below 1%. No statistically significant ancestry-related differences were observed in any gene, suggesting that, among early-onset FOLFOX-treated individuals, the TP53 pathway is similarly affected across H/L and NHW populations, with no evidence of ancestry-driven divergence in mutational profiles.

#### Early-Onset Non-FOLFOX Patients: Hispanic/Latino vs. Non-Hispanic White Comparisons

Table S9 compares the prevalence of TP53 pathway mutations between EOCRC H/L and NHW patients who did not receive FOLFOX treatment. TP53 alterations were the dominant event in both groups, occurring in 80.8% of H/L and 75.5% of NHW patients (p = 0.52), indicating similar mutation frequencies across ancestries. Alterations in ATM were more frequent among H/L patients (17.3%) compared with NHW patients (7.9%), a difference approaching borderline significance (p = 0.06). Mutations in CDKN2A and CHEK2 were rare (≤2%), while MDM2 and MDM4 mutations were absent in H/L patients and occurred in less than 1% of NHW cases.

#### Late-Onset FOLFOX-Treated Patients: Hispanic/Latino vs. Non-Hispanic White Comparisons

Table S10 presents the distribution of TP53 pathway gene mutations among LOCRC H/L and NHW patients treated with FOLFOX. TP53 mutations were highly prevalent and comparable across ancestries, observed in 73.4% of H/L and 75.5% of NHW patients (p = 0.53). Other pathway components showed low mutation frequencies, with no major ancestry-driven differences. Alterations in ATM, MDM2, MDM4, and CDKN2A occurred in less than 8% of cases overall and were similarly distributed between the two groups. However, CHEK2 mutations were slightly enriched in NHW patients (2.6%) compared with H/L patients (0.8%), a statistically significant difference (p = 0.022).

#### Late-Onset Non-FOLFOX Patients: Hispanic/Latino vs. Non-Hispanic White Comparisons

Table S11 compares TP53 pathway alterations between LOCRC H/L and NHW patients who did not receive FOLFOX therapy. TP53 mutations were common in both populations, identified in 72.0% of H/L and 68.1% of NHW patients (p = 0.68), indicating comparable mutation burdens across ancestries. Mutations in other pathway genes were infrequent and evenly distributed. ATM alterations, present in roughly 12% of both groups, represented the most recurrent secondary event. CDKN2A, CHEK2, MDM2, and MDM4 mutations occurred at low frequencies (<3%) and showed no statistically significant ancestry-related differences.

### 3.5 Mutational Landscape of the TP53 Pathway in Early-Onset H/L CRC

#### Early-Onset H/L CRC

Figure 1a illustrates the genomic landscape of TP53 pathway alterations in EOCRC H/L patients (n = 123), integrating mutation type, TMB, and FOLFOX treatment status. Overall, 88.6% of tumors carried at least one alteration within the TP53 pathway, underscoring its central role in H/L CRC biology. TP53 emerged as the dominant driver, exhibiting a high frequency of missense mutations (green) accompanied by truncating alterations, including frameshift deletions (light blue) and nonsense mutations (red). Secondary genes such as ATM and CDKN2A were mutated in smaller subsets (∼11% and 2%, respectively), primarily through loss-of-function variants. CHEK2 alterations were infrequent (∼2%), while MDM2 and MDM4 remained unaltered across the cohort. TMB levels were generally low, though a few outlier samples displayed elevated mutational burdens without a clear correlation to specific gene alterations. Cases treated with FOLFOX (blue) and untreated (red) were interspersed throughout the oncoplot, suggesting no distinct treatment-associated mutation pattern within the TP53 signaling network. Collectively, these findings highlight TP53 as the predominant mutational hotspot in early-onset H/L CRC, with co-alterations in DNA repair–related genes contributing modestly to pathway complexity.

**Figure 1.**
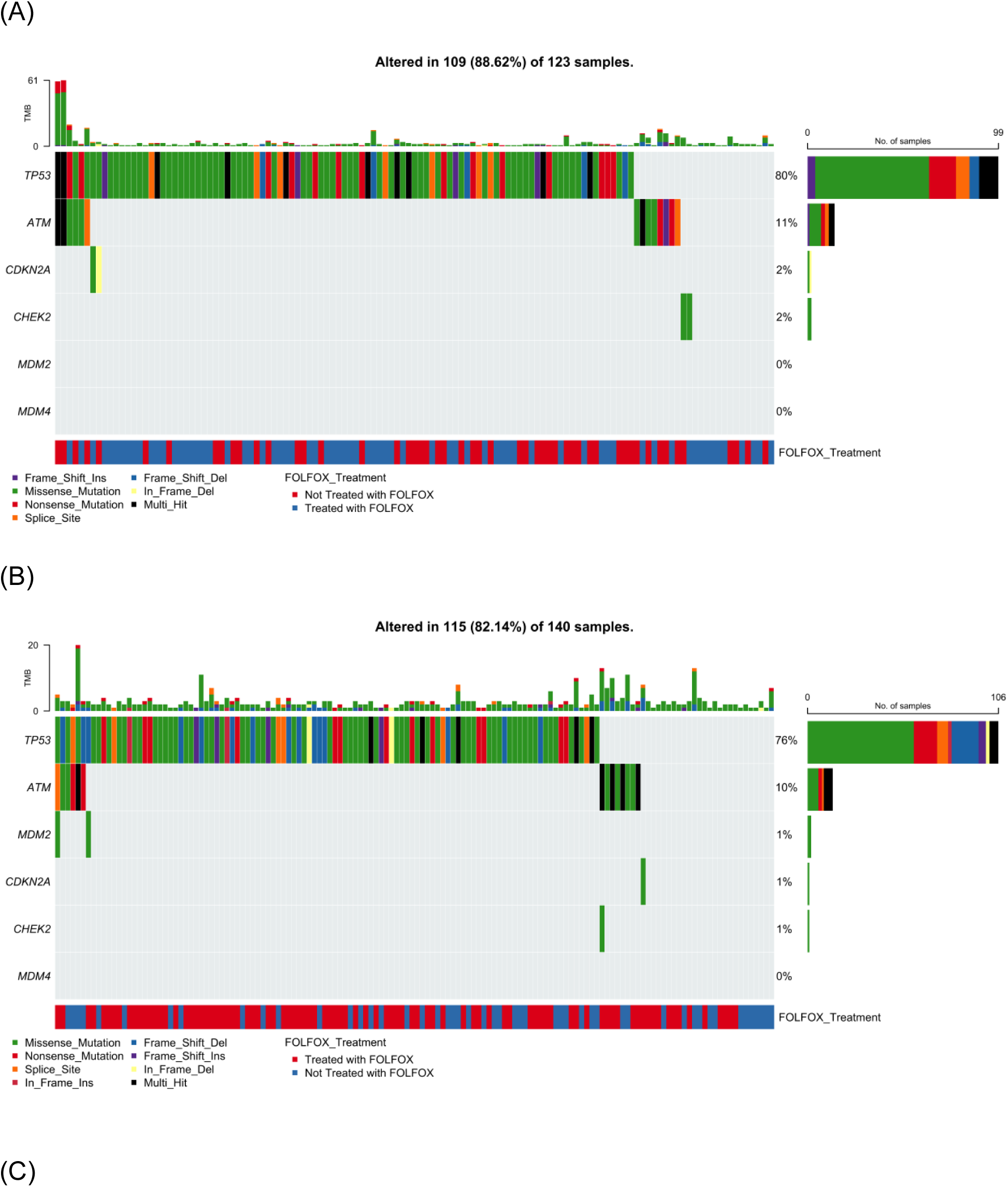
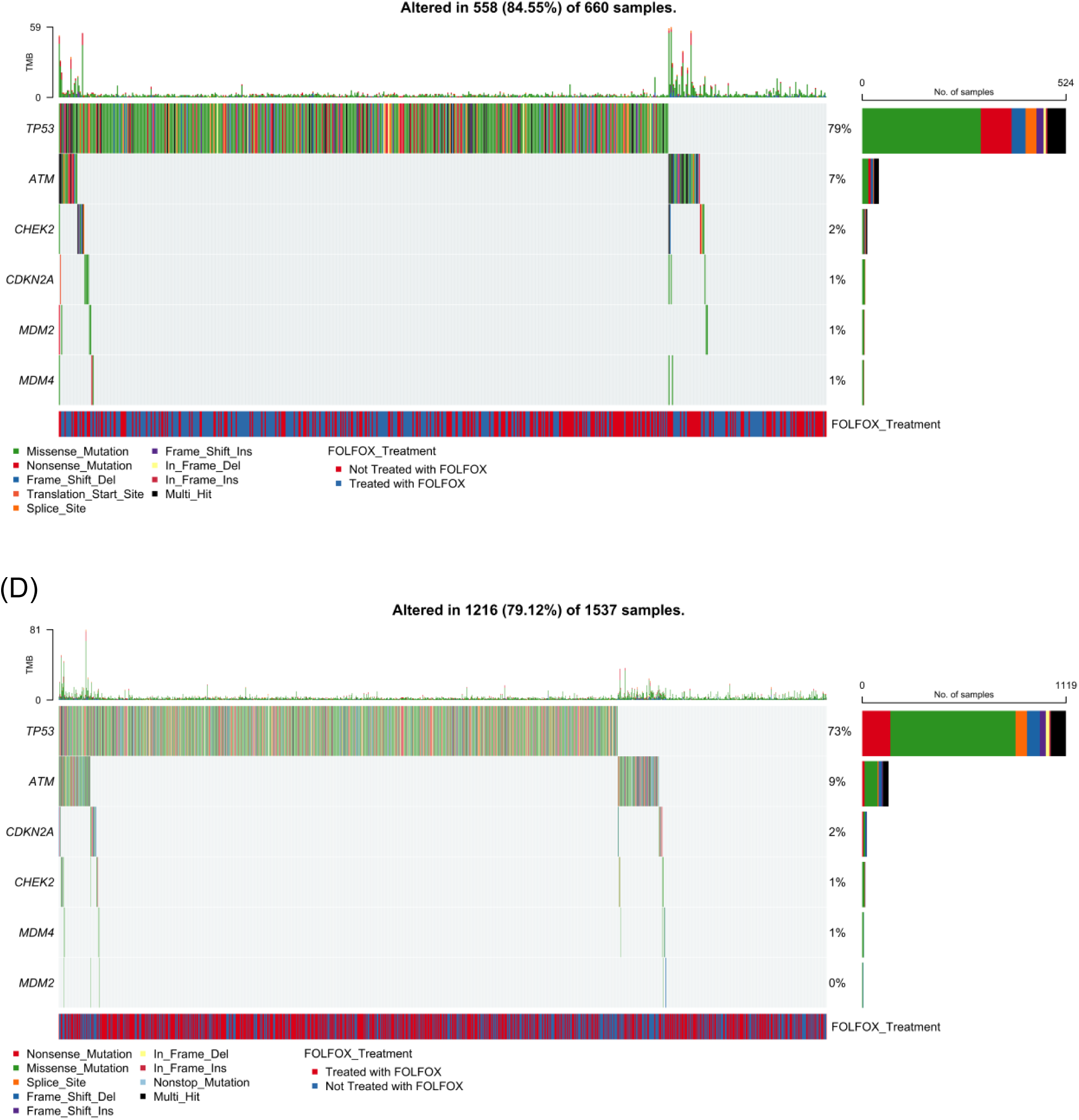
Somatic mutation landscape of TP53 pathway genes in colorectal cancer (CRC) stratified by age of onset and ancestry. Oncoplots showing gene-level mutation profiles of key TP53 pathway components in colorectal cancer, stratified by age of onset (early vs. late) and ancestry (Hispanic/Latino vs. Non-Hispanic White). Panels display mutation types, tumor mutational burden (TMB), and FOLFOX treatment status across: (A) early-onset Hispanic/Latino (H/L) patients, (B) late-onset H/L patients, (C) early-onset Non-Hispanic White (NHW) patients, and (D) late-onset NHW patients.

#### Late-Onset H/L CRC

Figure 1b illustrates the spectrum of TP53 pathway alterations in LOCRC H/L patients (n = 140), integrating mutation type, TMB, and FOLFOX treatment status. A total of 82.1% of tumors exhibited at least one somatic alteration within the TP53 pathway, emphasizing its persistent disruption in advanced-age disease. TP53 remained the dominant altered gene, displaying a rich diversity of mutation types including missense (green), frameshift (blue), and nonsense (red) variants, often occurring as multi-hit or truncating events. ATM was the second most frequently affected gene, harboring both missense and multi-hit mutations, while CDKN2A, MDM2, and CHEK2 showed low-frequency events (<2%). MDM4 alterations were absent in this cohort. TMB levels were predominantly low, though several samples exhibited localized hypermutation without clear gene-specific clustering. The distribution of FOLFOX-treated (blue) and non-treated (red) cases appeared intermixed across the mutational landscape, suggesting that treatment exposure did not strongly influence mutation type or frequency. Overall, these data reveal that TP53 pathway dysregulation remains a defining molecular feature in late-onset H/L CRC, with ATM serving as a recurrent secondary event.

#### Early-Onset NHW CRC

Figure 1c presents the TP53 pathway mutational spectrum in EOCRC NHW patients (n = 660), incorporating mutation type, TMB, and FOLFOX treatment status. Overall, 84.6% of tumors displayed at least one alteration within the TP53 signaling axis, confirming its pervasive disruption in early-onset disease. TP53 was the most frequently altered gene, with a predominance of missense mutations (green) interspersed with frameshift deletions (light blue), nonsense mutations (red), and occasional multi-hit events (black). Secondary alterations in ATM were relatively common, characterized by truncating and missense variants, whereas mutations in CDKN2A and CHEK2 appeared at much lower frequencies (≤2%). MDM2 and MDM4 alterations were exceedingly rare, detected in only isolated cases. TMB values remained largely low across the cohort, although a small subset of hypermutated tumors was observed without clear enrichment for any specific gene. Both FOLFOX-treated (blue) and non-treated (red) patients were evenly distributed along the mutation spectrum, suggesting that chemotherapy exposure did not influence the mutational pattern of TP53 or its pathway components. Collectively, these data emphasize TP53 as the principal genomic driver in early-onset NHW CRC, with limited contribution from other pathway regulators.

#### Late-Onset NHW CRC

Figure 1d depicts the TP53 pathway mutation landscape in LOCRC NHW patients (n = 1,537), integrating mutation type, TMB, and FOLFOX treatment status. In total, 79.1% of tumors exhibited at least one alteration in TP53 pathway genes, underscoring the pathway’s pervasive disruption in late-onset disease. TP53 remained the most frequently mutated gene, showing a predominance of missense mutations (green) with additional frameshift (blue), nonsense (red), and multi-hit (black) variants. ATM represented the second most altered gene, marked by recurrent loss-of-function and truncating events, while CDKN2A and CHEK2 mutations occurred at low but notable frequencies (∼2–3%). Alterations in MDM2 and MDM4 were rare, appearing in less than 1% of cases. TMB values were predominantly low across the cohort, though occasional hypermutated outliers were observed. The distribution of FOLFOX-treated (blue) and non-treated (red) samples was evenly spread, suggesting that chemotherapy exposure did not significantly influence mutation patterns. Altogether, these results highlight TP53 as the central genomic driver in late-onset NHW CRC, with ATM and CDKN2A contributing modestly to additional genomic instability within the pathway.

### 3.6 Prognostic Influence of TP53 Pathway Alterations by Age, Ancestry, and Treatment Status

To assess the prognostic significance of TGF-beta pathway alterations in CRC, we performed Kaplan–Meier survival analyses stratified by patient age at diagnosis, ancestry group, and exposure to FOLFOX chemotherapy.

In EOCRC H/L patients treated with FOLFOX (Figure 2a), alterations in the TP53 pathway were associated with significantly poorer overall survival (p = 0.014). Individuals harboring pathway mutations exhibited a sharper decline in survival probability within the initial 40–50 months, after which the curve stabilized. In contrast, patients without TP53 pathway alterations maintained consistently high survival throughout the follow-up period. The wider confidence intervals observed among altered cases reflect a smaller subgroup size. Collectively, these findings suggest that TP53 pathway dysregulation may serve as an adverse prognostic factor among early-onset H/L patients receiving FOLFOX chemotherapy.

**Figure 2.**
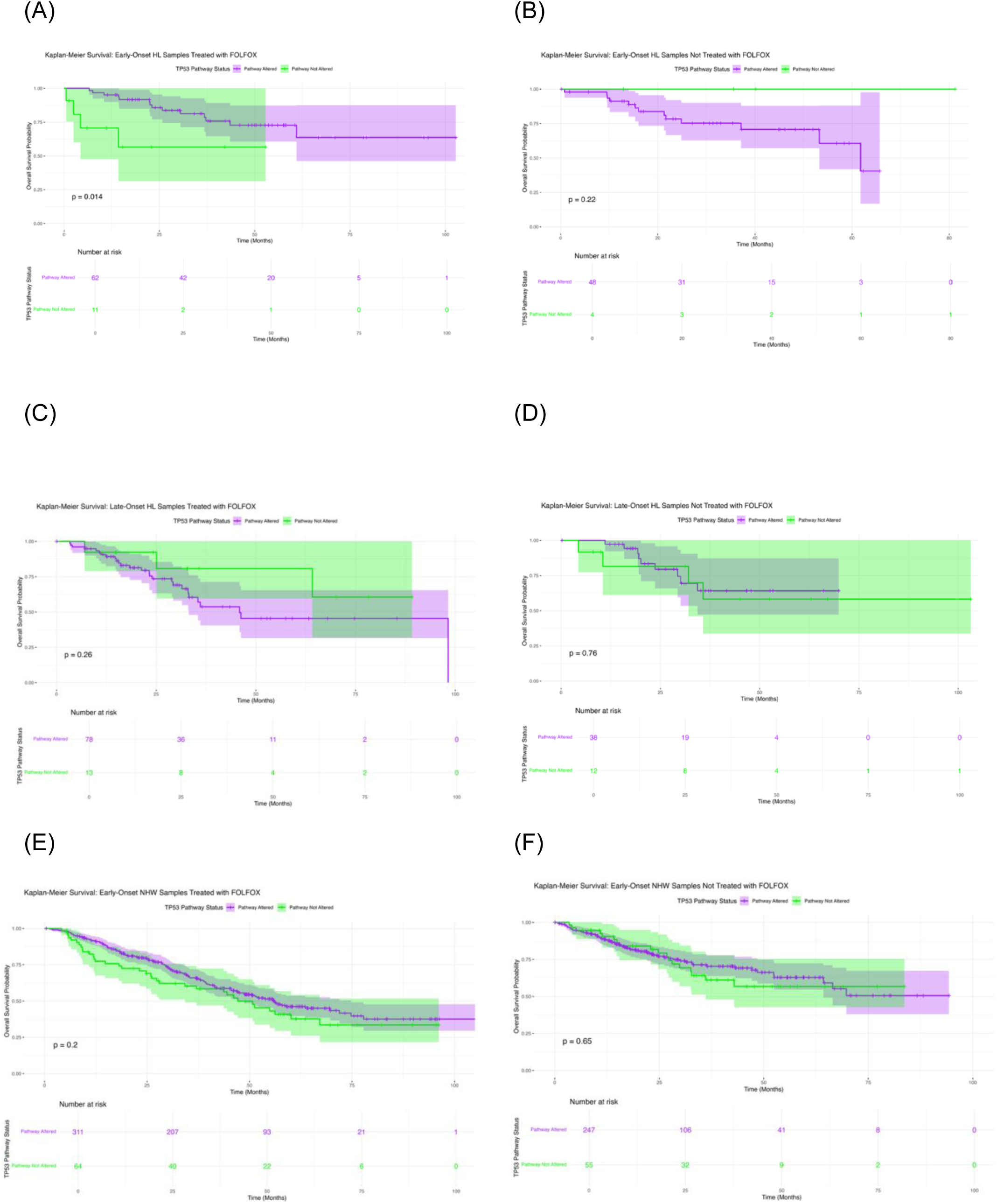
Kaplan–Meier analysis of overall survival by TP53 pathway alteration status across colorectal cancer (CRC) subgroups. This figure displays survival outcomes stratified by age of onset, ancestry, and FOLFOX treatment. Panels depict: (a) Early-Onset Hispanic/Latino (H/L) patients receiving FOLFOX, (b) Early-Onset H/L patients not treated with FOLFOX, (c) Late-Onset H/L patients treated with FOLFOX, (d) Late-Onset H/L patients not treated with FOLFOX, (e) Early-Onset Non-Hispanic White (NHW) patients receiving FOLFOX, and (f) Early-Onset NHW patients not receiving FOLFOX. Each curve compares cases harboring TP53 pathway alterations with those lacking such mutations. Shaded regions represent 95% confidence intervals, and accompanying tables show the number of patients at risk across selected time points.

In EOCRC H/L patients who did not undergo FOLFOX treatment (Figure 2b), overall survival did not significantly differ between those with and without TP53 pathway alterations (p = 0.22). Both subgroups demonstrated similarly favorable survival outcomes, with nearly parallel Kaplan–Meier curves throughout the follow-up period. Although a modest separation emerged toward the later months, this variation lacked statistical significance and may reflect the limited number of non-altered cases included in the analysis.

In LOCRC H/L patients who received FOLFOX chemotherapy (Figure 2c), overall survival did not differ significantly between those with TP53 pathway alterations and those without (p = 0.26). While patients lacking alterations appeared to maintain slightly higher survival probabilities early in follow-up, both curves aligned over time with broad, overlapping confidence intervals. These findings indicate that TP53 pathway mutation status had no clear impact on survival among late-onset H/L individuals treated with FOLFOX.

In LOCRC H/L patients who did not receive FOLFOX chemotherapy (Figure 2d), overall survival was comparable between those with and without TP53 pathway alterations (p = 0.76). The Kaplan–Meier curves for both groups closely overlapped across the entire observation period, with broad and intersecting confidence intervals indicating minimal divergence in outcomes. These findings suggest that TP53 pathway mutation status exerts little influence on survival in untreated late-onset H/L patients.

Among EOCRC NHW patients who received FOLFOX therapy (Figure 2e), overall survival did not differ significantly between those with and without TP53 pathway alterations (p = 0.20). The two survival trajectories remained largely parallel, with a modest divergence observed around mid-follow-up that did not reach statistical significance. Overlapping confidence intervals across most time points indicate that TP53 pathway status had no clear prognostic impact in this subgroup.

In EOCRC NHW patients who did not receive FOLFOX treatment (Figure 2f), overall survival did not significantly differ between those with TP53 pathway alterations and those without (p = 0.65). The Kaplan–Meier curves remained closely aligned throughout the follow-up period, with overlapping confidence intervals suggesting comparable survival probabilities across groups. These results indicate that TP53 pathway alterations do not appear to influence prognosis in untreated early-onset NHW cases.

In LOCRC NHW patients who received FOLFOX therapy (Figure S1a), TP53 pathway alteration status was not associated with significant differences in overall survival (p = 0.84). The Kaplan–Meier curves for altered and non-altered groups remained closely aligned throughout the follow-up, showing nearly identical survival probabilities and substantial overlap in confidence intervals. Both groups demonstrated gradual declines in survival over time, suggesting that TP53 pathway mutations do not meaningfully influence treatment response or prognosis in this FOLFOX-treated late-onset NHW cohort.

In LOCRC NHW patients who did not receive FOLFOX therapy (Figure S1b), overall survival did not significantly differ between individuals with TP53 pathway alterations and those without (p = 0.11). The Kaplan–Meier curves followed nearly parallel courses, with substantial overlap of their 95% confidence intervals across the study period. While a slight separation was observed beyond 60 months—suggesting a modest trend toward reduced survival among altered cases—this difference was not statistically significant, indicating that TP53 pathway alterations exert minimal influence on long-term outcomes in this untreated late-onset NHW cohort.

### 3.7 AI-assisted data exploration and pre-statistical discovery

The AI-HOPE and AI-HOPE-TP53 platforms were initially utilized to perform a focused, post-analytic interrogation of the integrated CRC datasets, enabling rapid generation of exploratory findings that informed downstream statistical analyses. Through natural language–based queries, the system identified multiple emerging patterns of potential biological and clinical relevance.

First, among EOCRC patients not treated with FOLFOX, AI-HOPE and AI-HOPE-TP53–driven exploratory analysis revealed multiple significant clinical and molecular differences between H/L and NHW groups. Using natural language–guided data interrogation, the platform identified key divergent attributes—including Ethnicity_Group, Treatment, Cancer Type, Race, MSI Type, TP53 mutation status, Sample Type, and Stage (Highest Recorded)—alongside continuous variables such as MSI Score, Diagnosis Age, and Current Age (Figure S2). These findings were confirmed through statistical validation (p < 0.05), highlighting distinct molecular and clinical patterns between EO H/L and NHW CRC patients. This AI-guided comparative profiling corresponds to the analyses presented in Section 3.6 (Figure 2a), providing foundational evidence of ancestry-related variation in CRC biology and supporting the prognostic relevance of TP53 pathway alterations within early-onset populations.

Second, AI-guided subgroup interrogation revealed a potential prognostic distinction linked to TP53 pathway alterations among EOCRC H/L patients treated with FOLFOX. Using the AI-HOPE and AI-HOPE-TP53 frameworks, the analysis compared the case cohort (EO H/L with TP53 pathway alterations, n = 62) against the control cohort (EO H/L without TP53 pathway alterations, n = 11). Subsequent Kaplan–Meier survival analysis confirmed that patients harboring TP53 pathway alterations exhibited significantly reduced overall survival compared with non-altered counterparts (log-rank p = 0.0139) (Figure S3). The survival trajectories demonstrated an early and sustained separation, with the altered group showing a steeper decline in survival probability over time. These results align with the broader survival trends reported in Section 3.6 (Figure 2b), suggesting that TP53 pathway alterations may represent a negative prognostic biomarker in early-onset H/L CRC patients receiving FOLFOX therapy and underscoring the clinical utility of AI-driven stratification in identifying high-risk molecular subgroups.

Third, AI-guided comparative analysis using the AI-HOPE-TP53 framework revealed a treatment-associated enrichment of TP53 mutations among LOCRC NHW patients. The analysis contrasted the case cohort of FOLFOX-treated individuals (n = 919) with the control cohort of untreated counterparts (n = 653), evaluating differences in mutation prevalence (Figure S4). TP53 mutations were identified in 73.45% of treated and 68.15% of untreated patients, representing a statistically significant distinction (p = 0.026; odds ratio = 1.293, 95% CI: 1.037–1.612). The modest but consistent elevation of TP53 mutations among treated cases suggests that FOLFOX exposure may be associated with a selection bias toward or persistence of TP53-altered tumors, potentially reflecting intrinsic resistance or altered treatment response dynamics. These findings emphasize the capacity of AI-driven interrogation to detect subtle, biologically meaningful treatment-related patterns, advancing understanding of molecular heterogeneity within late-onset CRC populations.

Finally, AI-guided interrogation using the AI-HOPE-TP53 framework revealed a treatment-associated difference in CDKN2A mutation frequency among LOCRC NHW patients. Comparative analysis between the FOLFOX-treated cohort (n = 919) and the untreated cohort (n = 653) demonstrated that CDKN2A mutations occurred in 0.98% of treated versus 2.45% of untreated patients (Figure S5). Statistical evaluation using Fisher’s exact test confirmed a significant reduction in CDKN2A mutation prevalence among the treated group (p = 0.036; odds ratio = 0.394, 95% CI: 0.173–0.897). This observation suggests that FOLFOX treatment may exert selective pressure against tumors harboring CDKN2A mutations or that such mutations are less compatible with treatment response. Collectively, these findings highlight the potential impact of therapy on the genomic architecture of late-onset CRC and underscore the utility of AI-based analytical frameworks in uncovering subtle, treatment-linked molecular variations within specific patient subgroups.

These preliminary AI-derived insights guided the identification of key subgroup comparisons for subsequent statistical evaluation. The AI-HOPE and AI-HOPE-TP53 platforms then autonomously performed data filtering and cohort generation by integrating clinical, molecular, and treatment variables, resulting in validated mutation frequency matrices and stratified survival outputs. This automated workflow minimized manual intervention, enhanced analytical reproducibility, and streamlined the progression from exploratory hypothesis discovery to rigorous confirmatory testing.

## 4. Discussion

In this multi-cohort analysis integrating 2,515 CRC across ancestry, age, and treatment strata, we used conventional statistics alongside two conversational AI systems (AI-HOPE and AI-HOPE-TP53) to interrogate how TP53-pathway alterations relate to clinicogenomic features and survival. Three findings stand out. First, TP53-pathway disruption is common across CRC, with missense variants predominating in every subgroup—consistent with the central role of TP53 in CRC biology. Second, we observed treatment-linked shifts in specific genes in late-onset (LO) NHW patients (e.g., enrichment of TP53 and depletion of ATM/CDKN2A among FOLFOX-treated cases), suggesting that chemotherapy exposure or treatment selection may shape the observable genomic landscape. Third—and most germane to precision oncology for disproportionately affected populations—TP53-pathway alterations in FOLFOX-treated early-onset (EO) H/L patients were associated with superior overall survival, an effect not seen in other ancestry-age-treatment combinations. Together, these results propose that the prognostic meaning of TP53-pathway alterations is not universal but contingent on clinical context, therapy, and population.

### Context within the literature

Prior studies emphasize TP53 as a pervasive driver in CRC and often link TP53 loss to adverse outcomes or chemoresistance across solid tumors. Yet CRC-specific evidence remains mixed, particularly regarding fluoropyrimidine- and platinum-based regimens. Our ancestry- and treatment-aware stratification adds nuance to this picture. The favorable association of TP53-pathway alterations with overall survival in FOLFOX-treated EO H/L patients contrasts with the commonly assumed negative prognostic role of TP53, indicating that (i) the functional consequences of TP53 variants may be context-dependent, and (ii) ancestry-linked biology, co-mutation patterns, tumor microenvironmental factors, or pharmacogenomic differences could modulate therapeutic effects of FOLFOX in ways not captured in largely NHW cohorts.

### Potential biological and clinical explanations

Several non–mutually exclusive mechanisms may underlie the finding observed in EOCRC H/L patients. One possible explanation involves the spectrum and function of TP53 variants. The relative proportion of missense, loss-of-function, and gain-of-function mutations may differ in EO H/L tumors, and certain missense variants could enhance sensitivity to DNA-damaging agents such as oxaliplatin and 5-fluorouracil, while others promote resistance. Another explanation lies in the co-alteration context. Therapy-associated differences in DNA damage response genes—such as ATM and CDKN2A— were noted in late-onset NHW cohorts. If EO H/L tumors with TP53-pathway alterations harbor fewer co-mutations in genes that drive chemoresistance or possess distinct cooperating alterations, including the TSC1 or RPTOR variants observed in non-FOLFOX-treated H/L tumors, their overall response to FOLFOX might be enhanced.

Tumor ecology and environmental exposure may also play important roles. Differences in age of onset and ancestry correlate with unique environmental influences, microbiome compositions, and immune profiles, any of which may modify how TP53-altered tumors respond to FOLFOX by affecting apoptosis or DNA repair capacity. Additionally, differences in treatment intensity and care delivery may contribute. Younger patients generally tolerate chemotherapy better, and if EO H/L patients are more likely to complete or maintain dose intensity, the cytotoxic effects of FOLFOX mediated through TP53-dependent pathways could be more fully realized.

These hypotheses suggest that the interaction between genetic, biological, and treatment-related factors shapes the observed prognostic association in EO H/L CRC. Functional validation using TP53-engineered experimental models that recapitulate the variant spectrum found in EO H/L tumors—coupled with oxaliplatin/5-FU response assays and studies incorporating microenvironmental perturbations—will be critical to elucidate the underlying mechanisms.

### Implications for precision oncology and health equity

Our findings argue against a “one-size-fits-all” interpretation of TP53 status in CRC prognosis. Instead, therapy- and ancestry-aware stratification appears essential. For EO H/L patients eligible for FOLFOX, TP53-pathway alteration could help refine prognostication and—pending validation—support treatment counseling. Conversely, the treatment-linked enrichment of TP53 and depletion of ATM/CDKN2A in LO NHW cohorts highlight how real-world therapy can bias the genomic snapshots we analyze; clinical sequencing data should be interpreted with treatment history front-of-mind.

More broadly, these results reinforce the need to (i) increase representation of H/L patients in genomic datasets and clinical trials, (ii) integrate ancestry-informed analyses prospectively, and (iii) examine pharmacogenomic variants and social determinants of health that could mediate access, adherence, dose intensity, and outcomes. Precision oncology that advances equity must couple molecular insights with context.

### Methodological advances: conversational AI as an engine for discovery

AI-HOPE and AI-HOPE-TP53 accelerated hypothesis generation by enabling natural-language queries that assembled cohorts, surfaced imbalances, and prioritized comparisons for formal testing. This human-in-the-loop loop—rapid exploratory AI followed by orthodox statistics (Fisher’s/χ², KM/log-rank, Cox)—reduced manual data wrangling, improved reproducibility, and exposed subtle patterns (e.g., therapy-linked shifts in ATM/CDKN2A) that might otherwise be overlooked. Importantly, AI outputs were treated as pre-statistical leads; only findings that survived conventional inference were highlighted. We view this workflow as a template for scalable, auditable, and equitable biomarker discovery.

### Limitations

Several limitations should be considered when interpreting these findings. First, the study’s retrospective and multi-source design introduces inherent variability in treatment documentation and timing. Despite applying stringent criteria to define FOLFOX exposure, residual misclassification cannot be fully excluded. Second, while Cox proportional hazards models were adjusted for age, sex, and ancestry, some datasets lacked uniform annotation of important covariates such as disease stage, microsatellite instability status, comorbidities, chemotherapy dose intensity, metastatic sites, and the use of biologic agents. The absence of these variables could introduce unmeasured confounding in the survival analyses. Third, certain early-onset strata—particularly those representing non-altered comparator groups—contained relatively small sample sizes, leading to wider confidence intervals and potential sensitivity to sampling variability.

Additionally, the definition of TP53 pathway alterations was restricted to nonsynonymous mutations in a curated set of genes. Future studies incorporating copy number changes, structural variants, transcriptomic activity scores, and pathway-level functional readouts may provide a more comprehensive understanding of TP53 dysregulation. Finally, although this investigation integrated multiple publicly available cohorts to enhance statistical power and diversity, the generalizability of the results remains limited. Prospective validation in larger, contemporary, and ethnically diverse populations— especially among EOCRC H/L patients—will be essential to confirm the prognostic and therapeutic implications of TP53 pathway alterations.

### Future directions

Prospective studies should (i) validate the favorable prognostic association of TP53-pathway alterations in FOLFOX-treated EO H/L CRC; (ii) parse TP53 variant classes and co-mutation signatures that best predict outcome; (iii) integrate transcriptional and functional readouts (e.g., p53 activity signatures, apoptosis priming, DNA repair capacity); (iv) incorporate pharmacogenomic loci relevant to 5-FU/oxaliplatin metabolism; and (v) couple molecular data with treatment delivery metrics and social determinants (35) to disentangle biology from access and adherence effects. Interventional trials could stratify by TP53-pathway status within EO H/L cohorts to test FOLFOX optimization or rational combinations (e.g., DNA damage response inhibitors) guided by co-alterations.

We demonstrate that the prognostic meaning of TP53-pathway alterations in CRC is context-specific. In FOLFOX-treated EOCRC H/L patients, TP53-pathway disruption associates with improved overall survival, while other ancestry-age-treatment strata show no benefit or therapy-linked genomic shifts without clear prognostic signal. By pairing conversational AI with rigorous statistics, we rapidly identified and validated these subgroup effects, illustrating how AI-enabled, equity-minded analytics can uncover clinically actionable patterns that traditional, ancestry-agnostic approaches miss. Prospective validation, multi-omic data integration (36) and mechanistic dissection are now warranted to translate these insights into tailored care for populations most impacted by EOCRC.

## Data Availability

All data used in the present study is publicly available at https://www.cbioportal.org/ and https://genie.cbioportal.org. The datasets used in our study were aggregated/summary data, and no individual-level data were used. Additional data can be provided upon reasonable request to the authors.

---

**Table S1.** Comparison of Early-Onset Hispanic/Latino (H/L) Patients Treated with FOLFOX versus Not Treated with FOLFOX.

| TP53 Pathway |  |  |  |
| --- | --- | --- | --- |
| Gene | Early-Onset Hispanic/Latino<br>Treated with FOLFOX<br>n (%) | Early-Onset Hispanic/Latino<br>Not Treated with FOLFOX<br>n (%) | p-value |
| TP53 Mutation |  |  |  |
| Present | 57 (78.1%) | 42 (80.8%) | 0.8876 |
| Absent | 16 (21.9%) | 10 (19.2%) |  |
| MDM2 Mutation |  |  |  |
| Present | 0 (0.0%) | 0 (0.0%) | 1 |
| Absent | 73 (100.0%) | 52 (100.0%) |  |
| MDM4 Mutation |  |  |  |
| Present | 0 (0.0%) | 0 (0.0%) | 1 |
| Absent | 73 (100.0%) | 52 (100.0%) |  |
| CDKN2A Mutation |  |  |  |
| Present | 1 (1.4%) | 1 (1.9%) | 1 |
| Absent | 72 (98.6%) | 51 (98.1%) |  |
| ATM Mutation |  |  |  |
| Present | 5 (6.8%) | 9 (17.3%) | 0.1236 |
| Absent | 68 (93.2%) | 43 (82.7%) |  |
| CHEK2 Mutation |  |  |  |
| Present | 1 (1.4%) | 1 (1.9%) | 1 |
| Absent | 72 (98.6%) | 51 (98.1%) |  |

**Table S2.** Comparison of Late-Onset Hispanic/Latino (H/L) Patients Treated with FOLFOX versus Not Treated with FOLFOX.

| TP53 Pathway |  |  |  |
| --- | --- | --- | --- |
| Gene | Late-Onset Hispanic/Latino<br>Treated with FOLFOX<br>n (%) | Late-Onset Hispanic/Latino<br>Not Treated with FOLFOX<br>n (%) | p-value |
| TP53 Mutation |  |  |  |
| Present | 71 (78.0%) | 36 (72.0%) | 0.5526 |
| Absent | 20 (22.0%) | 14 (28.0%) |  |
| MDM2 Mutation |  |  |  |
| Present | 2 (2.2%) | 0 (0.0%) | 0.539 |
| Absent | 89 (97.8%) | 50 (100.0%) |  |
| MDM4 Mutation |  |  |  |
| Present | 0 (0.0%) | 0 (0.0%) | 1 |
| Absent | 91 (100.0%) | 50 (100.0%) |  |
| CDKN2A Mutation |  |  |  |
| Present | 1 (1.1%) | 0 (0.0%) | 1 |
| Absent | 90 (98.9%) | 50 (100.0%) |  |
| ATM Mutation |  |  |  |
| Present | 8 (8.8%) | 6 (12.0%) | 0.7526 |
| Absent | 83 (91.2%) | 44 (88.0%) |  |
| CHEK2 Mutation |  |  |  |
| Present | 1 (1.1%) | 0 (0.0%) | 1 |
| Absent | 90 (98.9%) | 50 (100.0%) |  |

**Table S3.** Comparison of Early-Onset Non-Hispanic White (NHW) Patients Treated with FOLFOX versus Not Treated with FOLFOX.

| TP53 Pathway |  |  |  |
| --- | --- | --- | --- |
| Gene | Early-Onset NHW<br>Treated with FOLFOX<br>n (%) | Early-Onset NHW<br>Not Treated with FOLFOX<br>n (%) | p-value |
| TP53 Mutation |  |  |  |
| Present | 296 (78.9%) | 228 (75.5%) | 0.3319 |
| Absent | 79 (21.1%) | 74 (24.5%) |  |
| MDM2 Mutation |  |  |  |
| Present | 4 (1.1%) | 2 (0.7%) | 0.697 |
| Absent | 371 (98.9%) | 300 (99.3%) |  |
| MDM4 Mutation |  |  |  |
| Present | 1 (0.3%) | 4 (1.3%) | 0.178 |
| Absent | 374 (99.7%) | 298 (98.7%) |  |
| CDKN2A Mutation |  |  |  |
| Present | 3 (0.8%) | 5 (1.7%) | 0.4772 |
| Absent | 372 (99.2%) | 297 (98.3%) |  |
| ATM Mutation |  |  |  |
| Present | 19 (5.1%) | 24 (7.9%) | 0.171 |
| Absent | 356 (94.9%) | 278 (92.1%) |  |
| CHEK2 Mutation |  |  |  |
| Present | 5 (1.3%) | 8 (2.6%) | 0.3379 |
| Absent | 370 (98.7%) | 294 (97.4%) |  |

**Table S4.** Comparison of Late-Onset Non-Hispanic White (NHW) Patients Treated with FOLFOX versus Not Treated with FOLFOX.

| TP53 Pathway |  |  |  |
| --- | --- | --- | --- |
| Gene | Late-Onset NHW<br>Treated with FOLFOX<br>n (%) | Late-Onset NHW<br>Not Treated with FOLFOX<br>n (%) | p-value |
| TP53 Mutation |  |  |  |
| Present | 675 (73.4%) | 445 (68.1%) | 0.02559 |
| Absent | 244 (26.6%) | 208 (31.9%) |  |
| MDM2 Mutation |  |  |  |
| Present | 2 (0.2%) | 4 (0.6%) | 0.2404 |
| Absent | 917 (99.8%) | 649 (99.4%) |  |
| MDM4 Mutation |  |  |  |
| Present | 4 (0.4%) | 5 (0.8%) | 0.5023 |
| Absent | 915 (99.6%) | 648 (99.2%) |  |
| CDKN2A Mutation |  |  |  |
| Present | 9 (1.0%) | 16 (2.5%) | 0.03638 |
| Absent | 910 (99.0%) | 637 (97.5%) |  |
| ATM Mutation |  |  |  |
| Present | 66 (7.2%) | 79 (12.1%) | 0.001233 |
| Absent | 853 (92.8%) | 574 (87.9%) |  |
| CHEK2 Mutation |  |  |  |
| Present | 7 (0.8%) | 10 (1.5%) | 0.2276 |
| Absent | 912 (99.2%) | 643 (98.5%) |  |

**Table S5.** Comparison of Early-Onset versus Late-Onset Hispanic/Latino (H/L) Patients Treated with FOLFOX.

| TP53 Pathway |  |  |  |
| --- | --- | --- | --- |
| Gene | Early-Onset Hispanic/Latino<br>Treated with FOLFOX<br>n (%) | Late-Onset Hispanic/Latino<br>Treated with FOLFOX<br>n (%) | p-value |
| TP53 Mutation |  |  |  |
| Present | 57 (78.1%) | 71 (78.0%) | 1 |
| Absent | 16 (21.9%) | 20 (22.0%) |  |
| MDM2 Mutation |  |  |  |
| Present | 0 (0.0%) | 2 (2.2%) | 0.503 |
| Absent | 73 (100.0%) | 89 (97.8%) |  |
| MDM4 Mutation |  |  |  |
| Present | 0 (0.0%) | 0 (0.0%) | 1 |
| Absent | 73 (100.0%) | 91 (100.0%) |  |
| CDKN2A Mutation |  |  |  |
| Present | 1 (1.4%) | 1 (1.1%) | 1 |
| Absent | 72 (98.6%) | 90 (98.9%) |  |
| ATM Mutation |  |  |  |
| Present | 5 (6.8%) | 8 (8.8%) | 0.8676 |
| Absent | 68 (93.2%) | 83 (91.2%) |  |
| CHEK2 Mutation |  |  |  |
| Present | 1 (1.4%) | 1 (1.1%) | 1 |
| Absent | 72 (98.6%) | 90 (98.9%) |  |

**Table S6.** Comparison of Early-Onset versus Late-Onset Hispanic/Latino (H/L) Patients Not Treated with FOLFOX.

| TP53 Pathway |  |  |  |
| --- | --- | --- | --- |
| Gene | Early-Onset Hispanic/Latino<br>Not Treated with FOLFOX<br>n (%) | Late-Onset Hispanic/Latino<br>Not Treated with FOLFOX<br>n (%) | p-value |
| TP53 Mutation |  |  |  |
| Present | 42 (80.8%) | 36 (72.0%) | 0.4178 |
| Absent | 10 (19.2%) | 14 (28.0%) |  |
| MDM2 Mutation |  |  |  |
| Present | 0 (0.0%) | 0 (0.0%) | 1 |
| Absent | 52 (100.0%) | 50 (100.0%) |  |
| MDM4 Mutation |  |  |  |
| Present | 0 (0.0%) | 0 (0.0%) | 1 |
| Absent | 52 (100.0%) | 50 (100.0%) |  |
| CDKN2A Mutation |  |  |  |
| Present | 1 (1.9%) | 0 (0.0%) | 1 |
| Absent | 51 (98.1%) | 50 (100.0%) |  |
| ATM Mutation |  |  |  |
| Present | 9 (17.3%) | 6 (12.0%) | 0.6334 |
| Absent | 43 (82.7%) | 44 (88.0%) |  |
| CHEK2 Mutation |  |  |  |
| Present | 1 (1.9%) | 0 (0.0%) | 1 |
| Absent | 51 (98.1%) | 50 (100.0%) |  |

**Table S7.** Comparison of Early-Onset versus Late-Onset Non-Hispanic White (NHW) Patients Treated with FOLFOX.

| TP53 Pathway |  |  |  |
| --- | --- | --- | --- |
| Gene | Early-Onset NHW<br>Treated with FOLFOX<br>n (%) | Late-Onset NHW<br>Treated with FOLFOX<br>n (%) | p-value |
| TP53 Mutation |  |  |  |
| Present | 296 (78.9%) | 675 (73.4%) | 0.04582 |
| Absent | 79 (21.1%) | 244 (26.6%) |  |
| MDM2 Mutation |  |  |  |
| Present | 4 (1.1%) | 2 (0.2%) | 0.06222 |
| Absent | 371 (98.9%) | 917 (99.8%) |  |
| MDM4 Mutation |  |  |  |
| Present | 1 (0.3%) | 4 (0.4%) | 1 |
| Absent | 374 (99.7%) | 915 (99.6%) |  |
| CDKN2A Mutation |  |  |  |
| Present | 3 (0.8%) | 9 (1.0%) | 1 |
| Absent | 372 (99.2%) | 910 (99.0%) |  |
| ATM Mutation |  |  |  |
| Present | 19 (5.1%) | 66 (7.2%) | 0.2042 |
| Absent | 356 (94.9%) | 853 (92.8%) |  |
| CHEK2 Mutation |  |  |  |
| Present | 5 (1.3%) | 7 (0.8%) | 0.5134 |
| Absent | 370 (98.7%) | 912 (99.2%) |  |

**Table S8.**
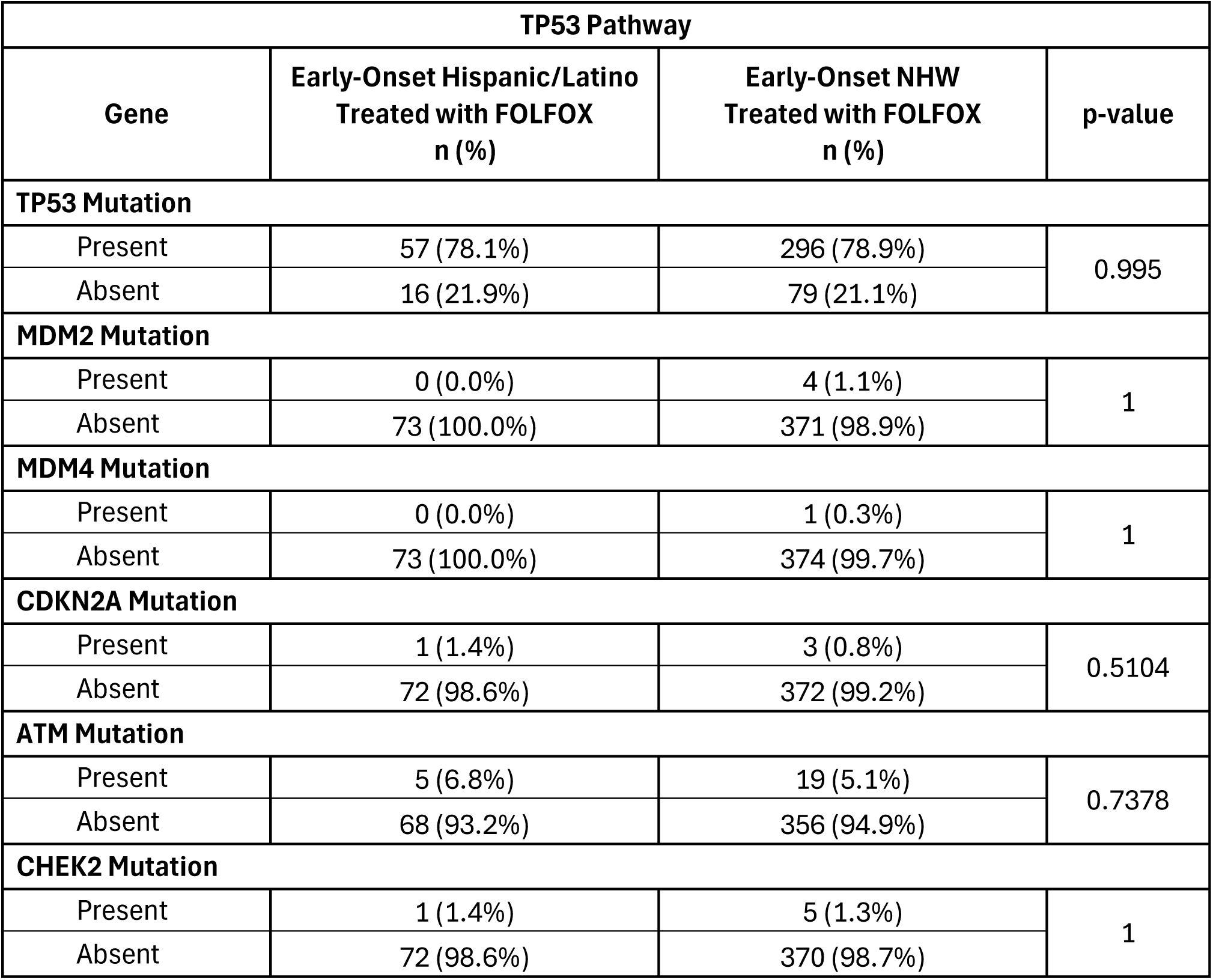
Comparison of Early-Onset Hispanic/Latino (H/L) versus Early-Onset Non-Hispanic White (NHW) Patients Treated with FOLFOX.

| TP53 Pathway |  |  |  |
| --- | --- | --- | --- |
| Gene | Early-Onset Hispanic/Latino<br>Treated with FOLFOX<br>n (%) | Early-Onset NHW<br>Treated with FOLFOX<br>n (%) | p-value |
| TP53 Mutation |  |  |  |
| Present | 57 (78.1%) | 296 (78.9%) | 0.995 |
| Absent | 16 (21.9%) | 79 (21.1%) |  |
| MDM2 Mutation |  |  |  |
| Present | 0 (0.0%) | 4 (1.1%) | 1 |
| Absent | 73 (100.0%) | 371 (98.9%) |  |
| MDM4 Mutation |  |  |  |
| Present | 0 (0.0%) | 1 (0.3%) | 1 |
| Absent | 73 (100.0%) | 374 (99.7%) |  |
| CDKN2A Mutation |  |  |  |
| Present | 1 (1.4%) | 3 (0.8%) | 0.5104 |
| Absent | 72 (98.6%) | 372 (99.2%) |  |
| ATM Mutation |  |  |  |
| Present | 5 (6.8%) | 19 (5.1%) | 0.7378 |
| Absent | 68 (93.2%) | 356 (94.9%) |  |
| CHEK2 Mutation |  |  |  |
| Present | 1 (1.4%) | 5 (1.3%) | 1 |
| Absent | 72 (98.6%) | 370 (98.7%) |  |

**Table S9.** Comparison of Early-Onset Hispanic/Latino (H/L) versus Early-Onset Non-Hispanic White (NHW) Patients Not Treated with FOLFOX.

| TP53 Pathway |  |  |  |
| --- | --- | --- | --- |
| Gene | Early-Onset Hispanic/Latino<br>Not Treated with FOLFOX<br>n (%) | Early-Onset NHW<br>Not Treated with FOLFOX<br>n (%) | p-value |
| TP53 Mutation |  |  |  |
| Present | 42 (80.8%) | 228 (75.5%) | 0.5163 |
| Absent | 10 (19.2%) | 74 (24.5%) |  |
| MDM2 Mutation |  |  |  |
| Present | 0 (0.0%) | 2 (0.7%) | 1 |
| Absent | 52 (100.0%) | 300 (99.3%) |  |
| MDM4 Mutation |  |  |  |
| Present | 0 (0.0%) | 4 (1.3%) | 1 |
| Absent | 52 (100.0%) | 298 (98.7%) |  |
| CDKN2A Mutation |  |  |  |
| Present | 1 (1.9%) | 5 (1.7%) | 1 |
| Absent | 51 (98.1%) | 297 (98.3%) |  |
| ATM Mutation |  |  |  |
| Present | 9 (17.3%) | 24 (7.9%) | 0.05927 |
| Absent | 43 (82.7%) | 278 (92.1%) |  |
| CHEK2 Mutation |  |  |  |
| Present | 1 (1.9%) | 8 (2.6%) | 1 |
| Absent | 51 (98.1%) | 294 (97.4%) |  |

**Table S10.** Comparison of Late-Onset Hispanic/Latino (H/L) versus Late-Onset Non-Hispanic White (NHW) Patients Treated with FOLFOX.

| TP53 Pathway |  |  |  |
| --- | --- | --- | --- |
| Gene | Late-Onset Hispanic/Latino<br>Treated with FOLFOX<br>n (%) | Late-Onset NHW<br>Treated with FOLFOX<br>n (%) | p-value |
| TP53 Mutation |  |  |  |
| Present | 675 (73.4%) | 228 (75.5%) | 0.5302 |
| Absent | 244 (26.6%) | 74 (24.5%) |  |
| MDM2 Mutation |  |  |  |
| Present | 2 (0.2%) | 2 (0.7%) | 0.2571 |
| Absent | 917 (99.8%) | 300 (99.3%) |  |
| MDM4 Mutation |  |  |  |
| Present | 4 (0.4%) | 4 (1.3%) | 0.1095 |
| Absent | 915 (99.6%) | 298 (98.7%) |  |
| CDKN2A Mutation |  |  |  |
| Present | 9 (1.0%) | 5 (1.7%) | 0.5181 |
| Absent | 910 (99.0%) | 297 (98.3%) |  |
| ATM Mutation |  |  |  |
| Present | 66 (7.2%) | 24 (7.9%) | 0.753 |
| Absent | 853 (92.8%) | 278 (92.1%) |  |
| CHEK2 Mutation |  |  |  |
| Present | 7 (0.8%) | 8 (2.6%) | 0.02249 |
| Absent | 912 (99.2%) | 294 (97.4%) |  |

**Table S11.**
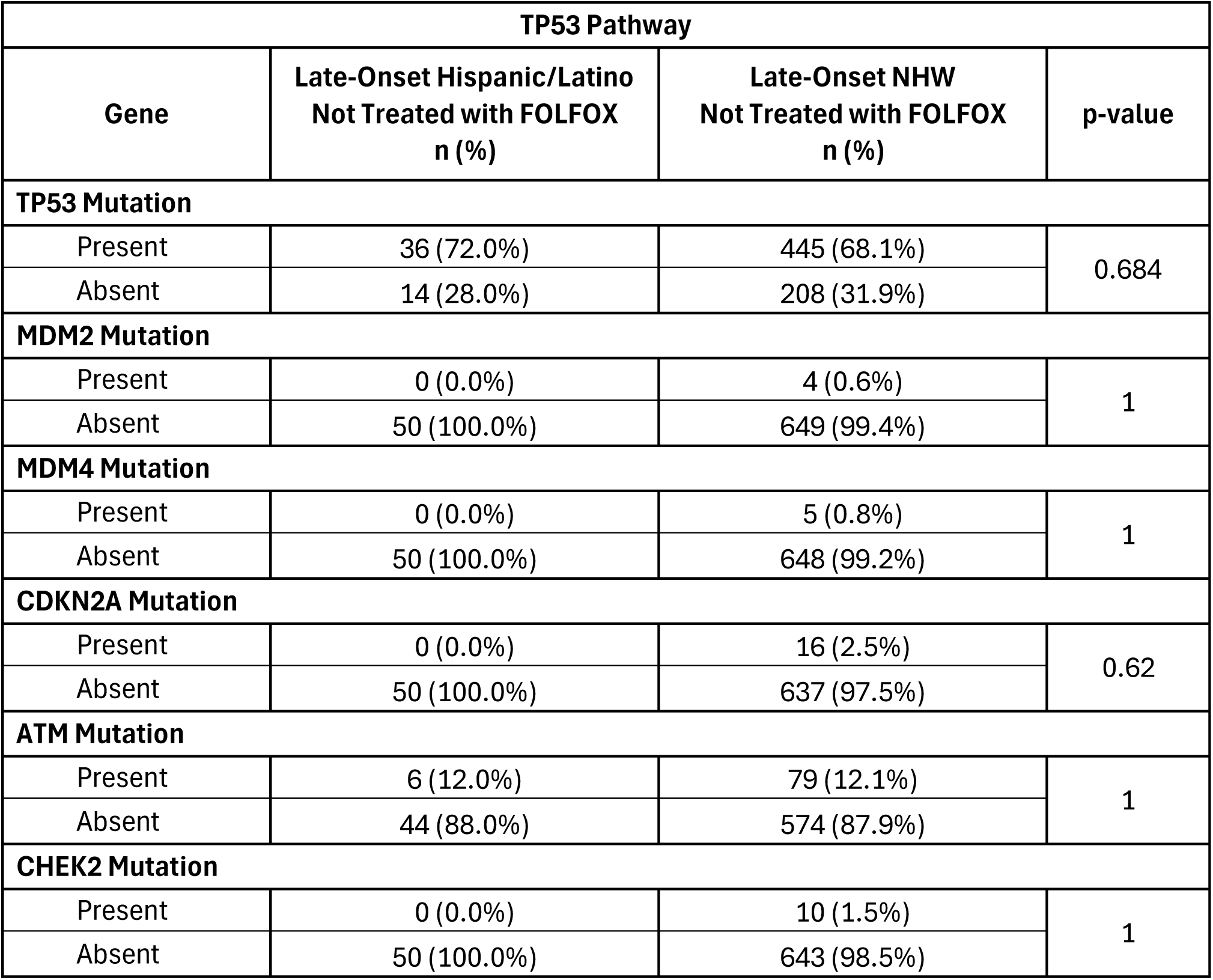
Comparison of Late-Onset Hispanic/Latino (H/L) versus Late-Onset Non-Hispanic White (NHW) Patients Not Treated with FOLFOX.

| TP53 Pathway |  |  |  |
| --- | --- | --- | --- |
| Gene | Late-Onset Hispanic/Latino<br>Not Treated with FOLFOX<br>n (%) | Late-Onset NHW<br>Not Treated with FOLFOX<br>n (%) | p-value |
| TP53 Mutation |  |  |  |
| Present | 36 (72.0%) | 445 (68.1%) | 0.684 |
| Absent | 14 (28.0%) | 208 (31.9%) |  |
| MDM2 Mutation |  |  |  |
| Present | 0 (0.0%) | 4 (0.6%) | 1 |
| Absent | 50 (100.0%) | 649 (99.4%) |  |
| MDM4 Mutation |  |  |  |
| Present | 0 (0.0%) | 5 (0.8%) | 1 |
| Absent | 50 (100.0%) | 648 (99.2%) |  |
| CDKN2A Mutation |  |  |  |
| Present | 0 (0.0%) | 16 (2.5%) | 0.62 |
| Absent | 50 (100.0%) | 637 (97.5%) |  |
| ATM Mutation |  |  |  |
| Present | 6 (12.0%) | 79 (12.1%) | 1 |
| Absent | 44 (88.0%) | 574 (87.9%) |  |
| CHEK2 Mutation |  |  |  |
| Present | 0 (0.0%) | 10 (1.5%) | 1 |
| Absent | 50 (100.0%) | 643 (98.5%) |  |

**Table S12.** Distribution of TP53 Pathway Mutation Classes Across Ancestry, Age of Onset, and FOLFOX Treatment in Colorectal Cancer. This table summarizes the proportional distribution of mutation types observed in six key genes of the TP53 signaling pathway, stratified by ancestry [Hispanic/Latino (H/L) vs. Non-Hispanic White (NHW)], disease onset category [early-onset (EO) vs. late-onset (LO)], and FOLFOX chemotherapy status (treated vs. untreated). Mutation classes encompass frame-shift insertions and deletions, in-frame insertions and deletions, missense and nonsense variants, nonstop mutations, splice site and splice region alterations, as well as translation start site changes. The percentages represent the relative contribution of each mutation class within a gene for the corresponding subgroup. Collectively, these data provide insight into the mutational spectrum and allow direct comparison of variant type distributions across ancestry, age, and treatment-defined patient populations.

|  | Hispanic/Latino Samples |  |  |  | Non-Hispanic White Samples |  |  |  |
| --- | --- | --- | --- | --- | --- | --- | --- | --- |
|  | Early-Onset |  | Late-Onset |  | Early-Onset |  | Late-Onset |  |
|  | Treated with FOLFOX | Not Treated with FOLFOX | Treated with FOLFOX | Not Treated with FOLFOX | Treated with FOLFOX | Not Treated with FOLFOX | Treated with FOLFOX | Not Treated with FOLFOX |
| **ATM** |  |  |  |  |  |  |  |  |
| Frame Shift Deletion | 16.7% | 0.0% | 18.2% | 12.5% | 10.0% | 8.9% | 11.0% | 12.3% |
| Frame Shift Insertion | 0.0% | 5.3% | 18.2% | 0.0% | 3.3% | 2.2% | 1.2% | 11.4% |
| In Frame Deletion | 0.0% | 0.0% | 0.0% | 0.0% | 0.0% | 0.0% | 1.2% | 0.0% |
| Missense Mutation | 66.7% | 47.4% | 54.5% | 62.5% | 46.7% | 75.6% | 61.0% | 62.3% |
| Nonsense Mutation | 16.7% | 36.8% | 0.0% | 25.0% | 36.7% | 11.1% | 15.9% | 12.3% |
| Splice Site | 0.0% | 10.5% | 9.1% | 0.0% | 3.3% | 2.2% | 9.8% | 1.8% |
| **CDKN2A** |  |  |  |  |  |  |  |  |
| Frame Shift Deletion | 0.0% | 0.0% | 0.0% | 0.0% | 0.0% | 0.0% | 20.0% | 31.3% |
| Frame Shift Insertion | 0.0% | 0.0% | 0.0% | 0.0% | 0.0% | 0.0% | 10.0% | 0.0% |
| In Frame Deletion | 0.0% | 100.0% | 0.0% | 0.0% | 0.0% | 0.0% | 0.0% | 0.0% |
| Missense Mutation | 100.0% | 0.0% | 100.0% | 0.0% | 100.0% | 80.0% | 40.0% | 43.8% |
| Nonsense Mutation | 0.0% | 0.0% | 0.0% | 0.0% | 0.0% | 0.0% | 30.0% | 25.0% |
| Translation Start Site | 0.0% | 0.0% | 0.0% | 0.0% | 0.0% | 20.0% | 0.0% | 0.0% |
| **CHEK2** |  |  |  |  |  |  |  |  |
| Frame Shift Deletion | 0.0% | 0.0% | 0.0% | 0.0% | 16.7% | 10.0% | 0.0% | 0.0% |
| Frame Shift Insertion | 0.0% | 0.0% | 0.0% | 0.0% | 33.3% | 0.0% | 28.6% | 0.0% |
| Missense Mutation | 100.0% | 100.0% | 100.0% | 0.0% | 33.3% | 60.0% | 71.4% | 80.0% |
| Nonsense Mutation | 0.0% | 0.0% | 0.0% | 0.0% | 0.0% | 20.0% | 0.0% | 0.0% |
| Splice Site | 0.0% | 0.0% | 0.0% | 0.0% | 16.7% | 10.0% | 0.0% | 20.0% |
| **MDM2** |  |  |  |  |  |  |  |  |
| Frame Shift Deletion | 0.0% | 0.0% | 0.0% | 0.0% | 0.0% | 0.0% | 100.0% | 0.0% |
| Missense Mutation | 0.0% | 0.0% | 100.0% | 0.0% | 100.0% | 50.0% | 0.0% | 100.0% |
| Nonsense Mutation | 0.0% | 0.0% | 0.0% | 0.0% | 0.0% | 50.0% | 0.0% | 0.0% |
| **MDM4** |  |  |  |  |  |  |  |  |
| Frame Shift Deletion | 0.0% | 0.0% | 0.0% | 0.0% | 0.0% | 0.0% | 0.0% | 20.0% |
| Missense Mutation | 0.0% | 0.0% | 0.0% | 0.0% | 0.0% | 100.0% | 100.0% | 80.0% |
| Nonsense Mutation | 0.0% | 0.0% | 0.0% | 0.0% | 100.0% | 0.0% | 0.0% | 0.0% |
| **TP53** |  |  |  |  |  |  |  |  |
| Frame Shift Deletion | 6.6% | 2.1% | 15.3% | 10.0% | 8.3% | 7.6% | 6.3% | 7.6% |
| Frame Shift Insertion | 3.3% | 6.3% | 4.2% | 2.5% | 4.6% | 3.6% | 3.8% | 2.3% |
| In Frame Deletion | 0.0% | 0.0% | 2.8% | 0.0% | 1.2% | 0.8% | 2.3% | 1.4% |
| In Frame Insertion | 0.0% | 0.0% | 2.8% | 0.0% | 0.6% | 0.8% | 0.5% | 0.8% |
| Missense Mutation | 63.9% | 68.8% | 54.2% | 72.5% | 64.4% | 64.0% | 65.9% | 67.8% |
| Nonsense Mutation | 19.7% | 14.6% | 15.3% | 7.5% | 16.0% | 16.8% | 15.3% | 13.1% |
| Nonstop Mutation | 0.0% | 0.0% | 0.0% | 0.0% | 0.0% | 0.0% | 0.1% | 0.0% |
| Splice Region | 0.0% | 0.0% | 0.0% | 2.5% | 0.3% | 0.0% | 0.1% | 0.0% |
| Splice Site | 6.6% | 8.3% | 5.6% | 5.0% | 4.6% | 6.4% | 5.5% | 7.0% |

**Figure S1.**
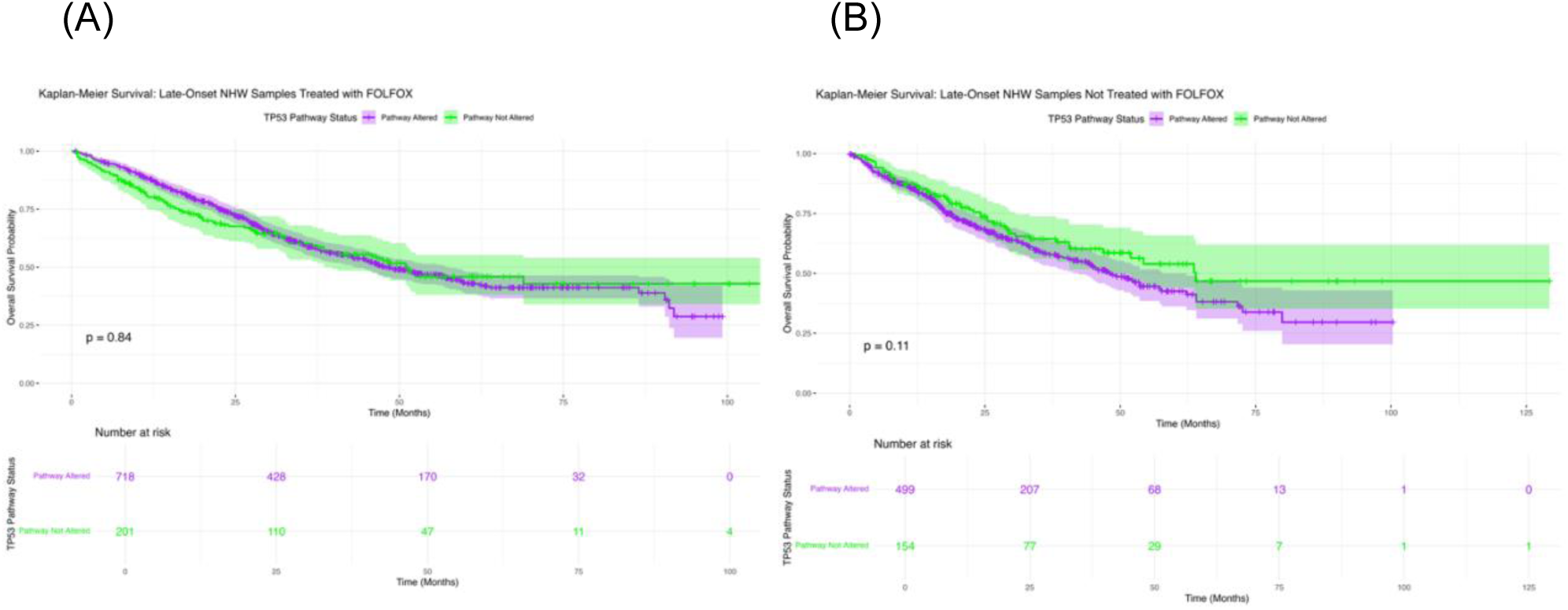
Kaplan–Meier analysis of overall survival by TP53 pathway alteration status in late-onset Non-Hispanic White (NHW) colorectal cancer (CRC) patients. Survival outcomes are stratified according to FOLFOX treatment status, with panels illustrating (a) late-onset NHW patients who received FOLFOX therapy and (b) late-onset NHW patients who did not. Each curve contrasts cases with TP53 pathway alterations against those without, highlighting potential treatment-related survival differences within this subgroup.

**Figure S2.**
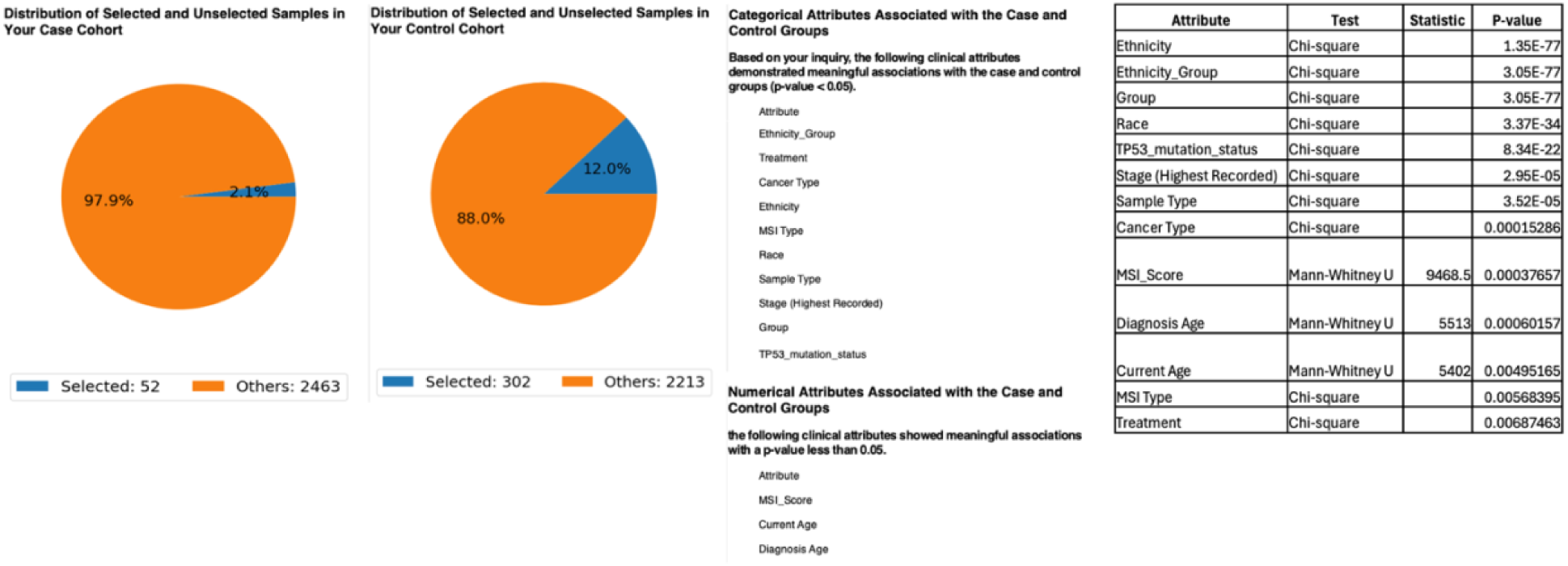
AI-HOPE-TP53-guided identification of significant clinical and molecular attributes distinguishing early-onset Hispanic/Latino (H/L) and Non-Hispanic White (NHW) colorectal cancer (CRC) patients not treated with FOLFOX. The figure summarizes the comparison between the case cohort (early-onset H/L, n = 52) and the control cohort (early-onset NHW, n = 302). Pie charts depict the proportion of selected versus unselected samples in each group. The accompanying tables list all categorical and numerical clinical attributes showing statistically significant differences between the cohorts (p < 0.05). Key categorical variables include Ethnicity_Group, Treatment, Cancer Type, Race, MSI Type, TP53 mutation status, Sample Type, and Stage (Highest Recorded). Significant continuous variables include MSI Score, Diagnosis Age, and Current Age. These AI-based analyses reveal multiple layers of divergence between early-onset Hispanic/Latino and Non-Hispanic White CRC patients, emphasizing distinct clinical and molecular profiles relevant to this study.

**Figure S3.**
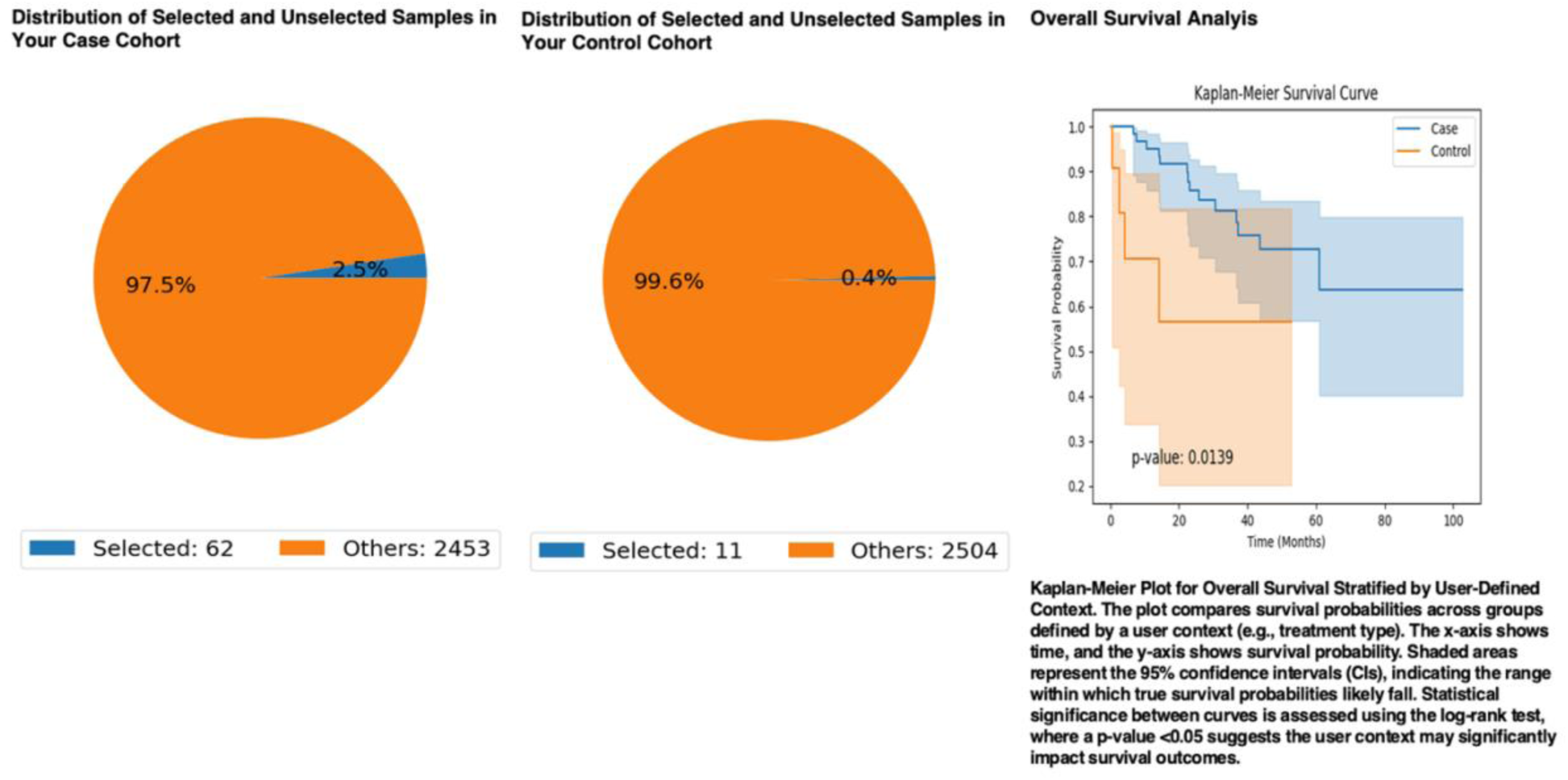
AI-guided selection and survival analysis of early-onset Hispanic/Latino (H/L) colorectal cancer (CRC) patients treated with FOLFOX, stratified by TP53 pathway alteration status. The AI-HOPE and AI-HOPE-TP53 platforms were utilized to define case and control cohorts based on integrative clinical, genomic, and treatment features. (Left) Pie charts illustrate the distribution of selected versus unselected samples for the case cohort—early-onset H/L CRC patients treated with FOLFOX and harboring TP53 pathway alterations (n = 62)—and the control cohort—early-onset H/L CRC patients treated with FOLFOX but without TP53 pathway alterations (n = 11). (Right) Kaplan–Meier overall survival (OS) analysis demonstrates that TP53 pathway–altered cases exhibited significantly reduced survival compared to non-altered controls (log-rank p = 0.0139). Shaded areas represent 95% confidence intervals. The observed separation of survival curves indicates a potential positive prognostic effect of TP53 pathway alterations among early-onset H/L patients receiving FOLFOX chemotherapy, supporting the results of this study.

**Figure S4.**
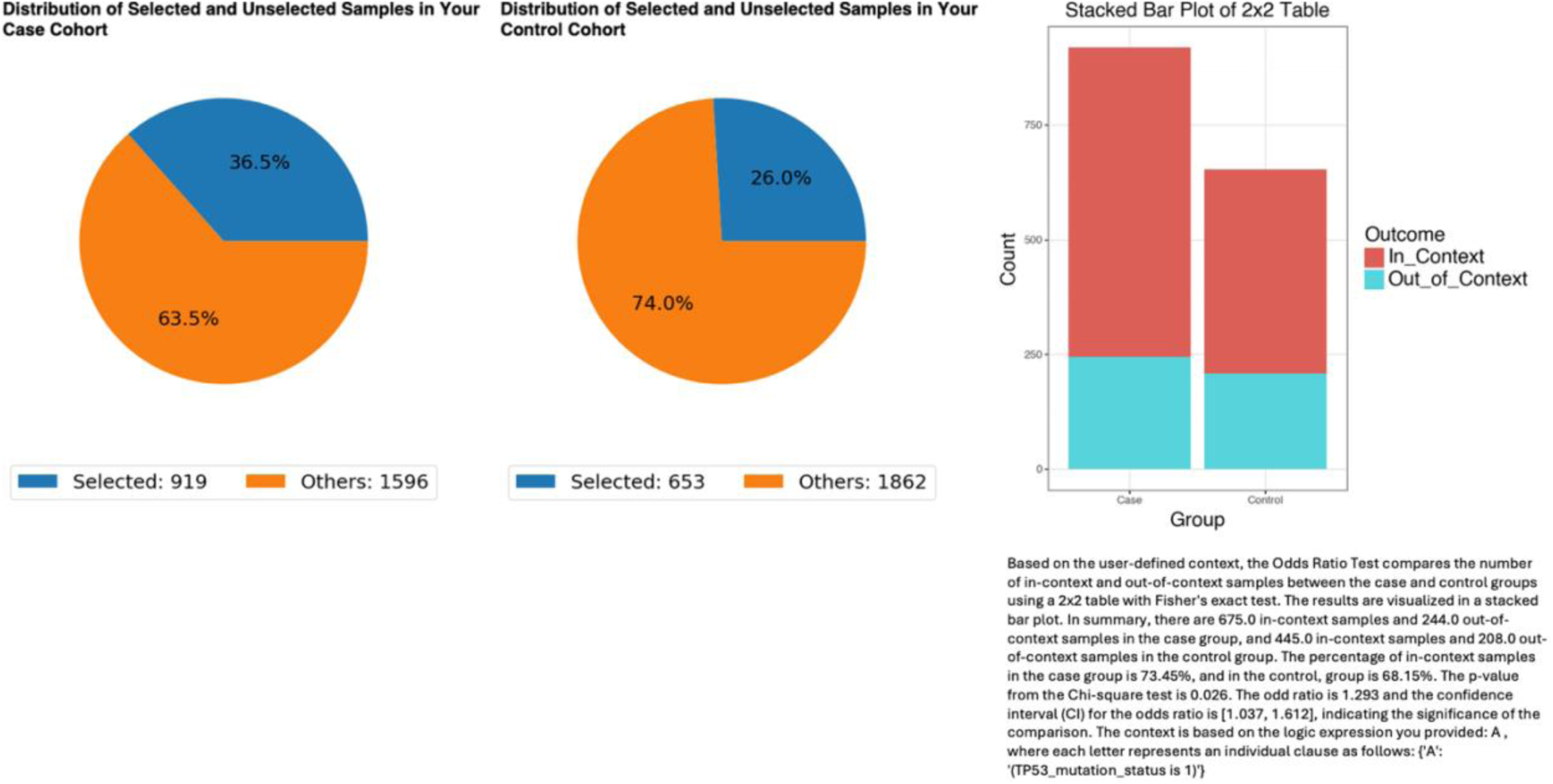
Comparison of TP53 mutation frequency between late-onset Non-Hispanic White (NHW) colorectal cancer (CRC) patients treated versus not treated with FOLFOX. The AI-HOPE and AI-HOPE-TP53 analytical frameworks were used to evaluate the prevalence of TP53 mutations in treatment-stratified late-onset NHW CRC cohorts. (Left) Pie charts illustrate the proportion of selected (in-context) versus unselected (out-of-context) samples in the case cohort—late-onset NHW CRC patients treated with FOLFOX (n = 919)—and the control cohort—late-onset NHW CRC patients not treated with FOLFOX (n = 653). (Right) A stacked bar plot displays the distribution of in-context and out-of-context samples across both groups, analyzed using Fisher’s exact test. TP53 mutations were detected in 73.45% of treated and 68.15% of untreated patients, yielding a p-value = 0.026 and an odds ratio = 1.293 (95% CI: 1.037–1.612), indicating a modest but statistically significant enrichment of TP53 mutations in the FOLFOX-treated cohort. These AI-derived findings suggest potential treatment-related selection or biological enrichment of TP53-altered tumors among late-onset NHW CRC patients, supporting the broader mechanistic insights described in this study.

**Figure S5.**
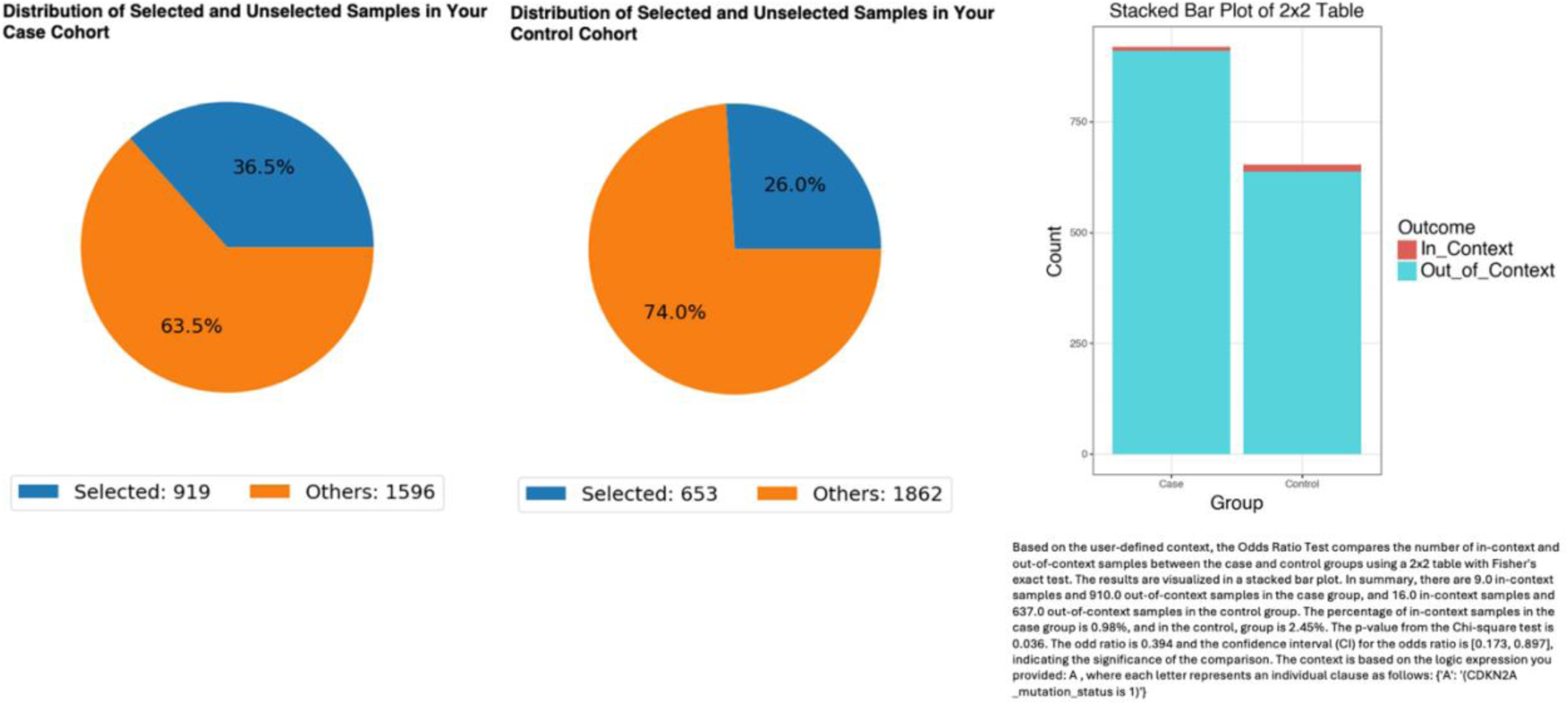
Comparison of CDKN2A mutation frequency between late-onset Non-Hispanic White (NHW) colorectal cancer (CRC) patients treated versus not treated with FOLFOX. The AI-HOPE and AI-HOPE-TP53 analytical frameworks were used to examine the prevalence of CDKN2A mutations among treatment-stratified late-onset NHW CRC cohorts. (Left) Pie charts depict the proportion of selected (in-context) versus unselected (out-of-context) samples in the case cohort—late-onset NHW CRC patients treated with FOLFOX (n = 919)—and the control cohort—late-onset NHW CRC patients not treated with FOLFOX (n = 653). (Right) A stacked bar plot summarizes the number of in-context and out-of-context samples across both groups, analyzed using Fisher’s exact test. CDKN2A mutations were observed in 0.98% of treated and 2.45% of untreated patients, yielding a p-value = 0.036 and an odds ratio = 0.394 (95% CI: 0.173–0.897), indicating a significantly lower frequency of CDKN2A mutations among patients treated with FOLFOX. These AI-guided results suggest potential treatment-associated genomic differences in tumor suppressor gene alterations within late-onset NHW CRC, providing additional molecular context for the findings presented in this study.

